# Prognostic pan-cancer and single-cancer models: A large-scale analysis using a real-world clinico-genomic database

**DOI:** 10.1101/2023.12.18.23300166

**Authors:** Sarah F. McGough, Svetlana Lyalina, Devin Incerti, Yunru Huang, Stefka Tyanova, Kieran Mace, Chris Harbron, Ryan Copping, Balasubramanian Narasimhan, Robert Tibshirani

## Abstract

Prognostic models in oncology have a profound impact on personalized cancer care and patient profiling, but tend to be heterogeneously developed and implemented in narrow patient cohorts. Here, we develop and benchmark multiple machine learning models to predict survival in pan-cancer and 16 single-cancer settings using a de-identified clinico-genomic database of 28,079 US patients with cancer. We identify key predictors of cancer prognosis, including 15 shared across seven or more cancer types, revealing strong consistency in cancer prognostic factors. We demonstrate that pan-cancer models generally outperform or match single-cancer models in predicting survival and risk stratifying patients, especially in smaller cancer cohorts, suggesting a unique transfer learning advantage of pan-cancer models. This work demonstrates the potential of pan-cancer approaches in enhancing the accuracy and applicability of prognostic models in oncology, paving the way for more personalized and effective cancer care strategies.

## Introduction

Prognostic models — models which predict a future health state, like survival — have a direct and important impact in precision oncology. In clinical practice, prognosis informs personalized treatment and care management by helping identify the future course of illness, appropriate course of therapy (i.e. ranging from aggressive treatment to surveillance), and resource allocation^1^. In clinical studies, stratifying patients into prognostic risk categories can aid in patient recruitment and trial enrichment for high-risk patients^2,3^. And critically, prognostic models have a profound impact on patient care, with these strategies enhancing quality of life and care by guiding clinicians towards the best treatment options tailored to each patient’s unique health profile. Typically, prognostic models are developed using a few disease-specific prognostic factors collected in routine clinical practice and used to predict patient survival or risk of death^4–6^.

The recent availability of large volumes of longitudinal, highly curated, and often linked patient-level health data from digital sources such as electronic health records (EHR) and genomic sequencing is contributing to advances in precision medicine by routinely collecting and storing millions of data points that offer a much more comprehensive patient profile. Machine learning models can learn from these high-dimensional datasets more effectively, bringing an opportunity for researchers to develop prognostic models that better leverage the myriad of prognostic factors from the patient’s health profile - not only illuminating key drivers in patient prognosis, but also driving improved and more personalized patient care.

Separately, an emerging paradigm in oncology is that of *pan-cancer* (cancer agnostic) research and treatment, in which cancer is characterized by genetic and molecular features rather than by its site of origin in the body. Indeed, multiple therapies have been approved in the last 5 years to treat a collection of cancer types on the basis of shared genetic mutations or predictive biomarkers that have been discovered to benefit from targeted treatment, such as tumor mutational burden (TMB)^7^ and fusions on the *NTRK* gene^8^. Pan-cancer prognostic factors –- particularly genomic or molecular in origin –- are another area of research that have shown early promise but warrant deeper exploration^9^. Further, whether pan-cancer settings provide a unique learning opportunity for prognostic models, over those typically developed in single cancer settings, is unknown^10^.

Although real-world prognostic models have been developed in the literature, a majority have been constructed using either clinical^9,11^ or genomic^12^ data alone, or within specific disease settings^13^. To advance our understanding of pan-cancer prognosis, it is essential to broaden the scope. Here we access a large, heterogeneous, and multi-cancer clinico-genomic database that offers a powerful tool for understanding both cancer genomics and clinical factors that impact survival under a pan-cancer paradigm. To our knowledge, the present study is the first large-scale analysis combining clinical and genomic data to evaluate and compare predictions in pan-cancer and dozens of single-cancer settings.

Our contributions are as follows: we systematically build and benchmark multiple pan-cancer and single-cancer machine learning prognostic models ranging in complexity using a large real-world clinico-genomic database; we identify key pan-cancer and single-cancer factors both shared and unique to each patient setting; and finally, on the basis of these factors, we risk stratify patients into prognostic subgroups. We compare the performance of pan- and single-cancer models to assess where pan-cancer models can provide advantage, and discuss implications for clinical and research settings.

## Results

### Pan- and single-cancer systematic modeling framework

We endeavored to create a systematic, reproducible framework for the building and benchmarking of multiple pan- and single-cancer prognostic models, outlined in Figure 1. This framework represents an end-to-end, data-driven process governing feature engineering, model building, model prediction, and model evaluation to enable comparisons between models and between cancer settings.

**Figure 1.**
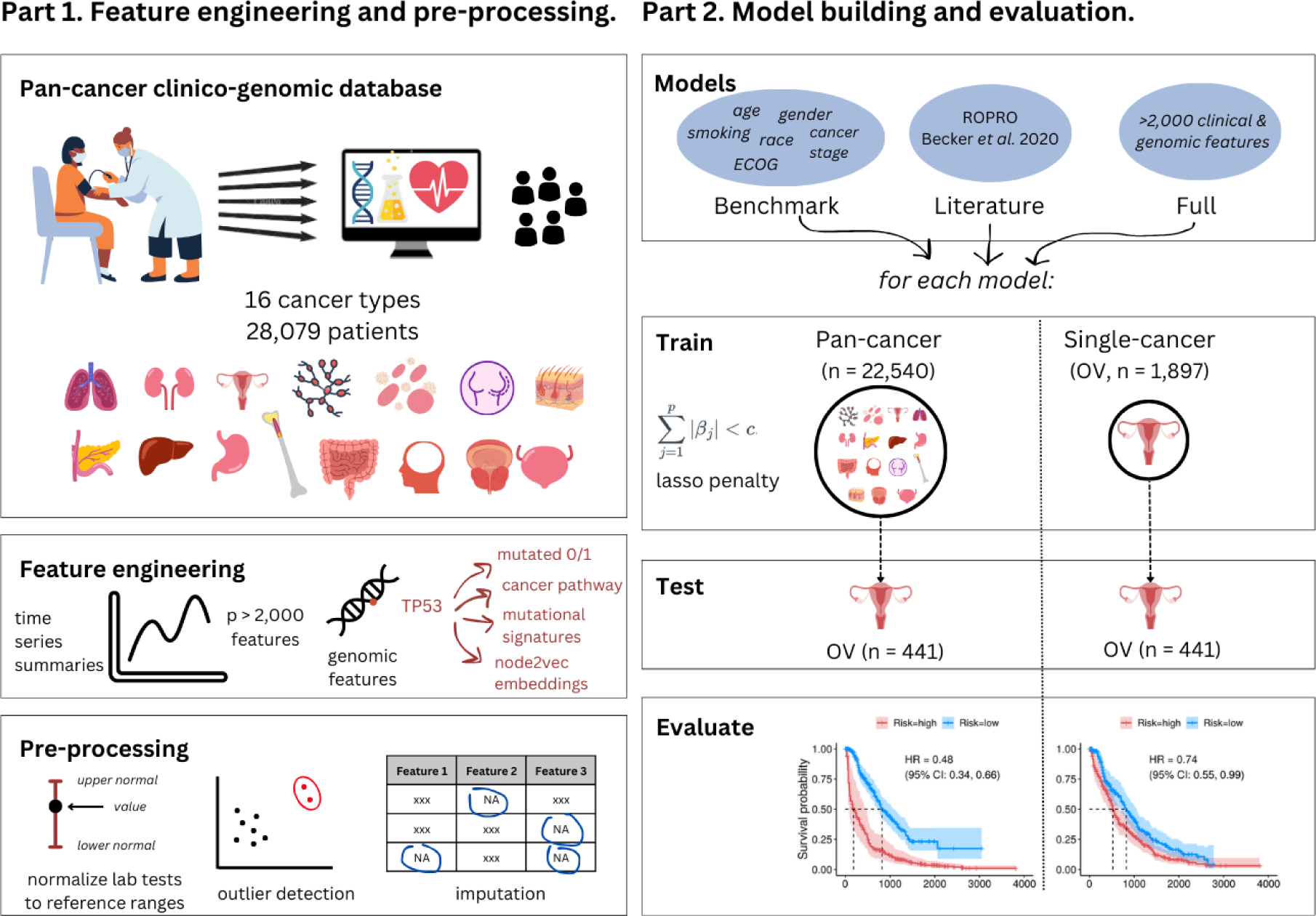
Pan- and single-cancer modeling framework. A US nationwide clinico-genomic database containing 28,079 patients and 16 cancer types was used to engineer over 2,000 features representing different modalities including demographics, treatment, laboratory tests and vital signs (represented as time series summaries), and genomics (represented as binary mutation status, affected cancer pathway, mutational signatures, and node2vec embeddings). Data were uniformly pre-processed including steps like outlier detection and processing and imputation. All steps are described in detail in *Materials & Methods* and are performed separately in train and test data where appropriate. Following this, multiple models were constructed with different feature sets: “benchmark” containing simple clinical features; “ROPRO” containing clinical features validated in the literature; “full” containing all 2,135 one-hot encoded clinico-genomic features. These models were trained both in pan-cancer and single-cancer cohorts, and evaluated in a single-cancer, out-of-sample test set for their ability to predict survival and risk stratify patients into high- and low-risk groups.

We obtained retrospective data on 28,079 patients from 16 different cancer cohorts with a recorded first line of therapy (1L) between January 1, 2011 and June 30, 2020 in a US clinico-genomic database (CGDB) linking longitudinal, patient-level electronic health records (EHRs) with patient-level tumor genomic profiling of >300 cancer-related genes^14,15^.

For each patient, we derived over 2,000 features representing 5 data modalities: clinical/demographic, laboratory/vital signs, treatment, cancer-specific (these 4 modalities collectively referred to as “clinical”), and genomic (Table S1). For each of the hundreds of individual lab tests in the database, we computed multiple time series summaries up until the point of prediction (1L initiation date). Genomic data, which contributed a majority of features, were used to characterize: (i) the alteration status (mutated or wild type) of >300 cancer-related genes for three variant types (short variant, copy number, and rearrangement), (ii) cancer biology pathways affected by these alterations, (iii) mutational signatures defined by the Catalogue Of Somatic Mutations In Cancer (COSMIC), and (iv) underlying protein interaction networks of affected genes (“node2vec”). In total, 2,059 features were derived (increasing to 2,135 model input features after one-hot encoding).

The pan-cancer cohort was highly heterogeneous with respect to cancer type and key clinico-genomic factors (Table 1). A majority of patients had solid tumor cancer diagnoses such as non-small cell lung cancer (NSCLC, n = 7,157, 25.4%), colorectal cancer (CRC, n = 5,059, 18.0%), and breast cancer (n = 4,801, 17.1%). Patient sample sizes for hematological (blood, bone marrow, and lymph node) diagnoses were considerably smaller, notably for diffuse large B-cell lymphoma (DLBCL, n = 163, 0.6%) and chronic lymphocytic leukemia (CLL, n = 109, 0.4%). Patient age ranged from 18 to 85 (median: 64 years; interquartile range, (IQR): 56, 72 years) and the median year of frontline therapy was 2017 (IQR: 2015, 2018). Approximately 42% of the pan-cancer cohort were never-smokers, though this ranged from 4% to 62% across individual cancer cohorts. Aligning with well-studied cancer biology, the TP53 gene short variant (SV), which encodes a tumor suppressor protein, was the most frequent alteration in the pan-cancer cohort with 62.5% of all patients having the alteration.

**Table 1.** Cohort characteristics.

|  | Pan cancer<br>(N=28,079) | Breast<br>(N=4,801) | CLL<br>(N=109) | Colorectal<br>(N=5,059) | DLBCL<br>(N=163) | Gastric<br>(N=1,712) | Head and Neck<br>(N=385) | atocellular Carcino<br>(N=183) | Melanoma<br>(N=717) | Multiple Myeloma<br>(N=325) | Non-Small Cell<br>(N=7,157) | Ovarian<br>(N=2,338) | Pancreatic<br>(N=1,918) | Prostate<br>(N=1,235) | Renal<br>(N=648) | Small Cell<br>(N=446) | Urothelial<br>(N=883) |
| --- | --- | --- | --- | --- | --- | --- | --- | --- | --- | --- | --- | --- | --- | --- | --- | --- | --- |
| **Age** |  |  |  |  |  |  |  |  |  |  |  |  |  |  |  |  |  |
| Mean (SD) | 63.2 (11.8) | 58.1 (12.2) | 65.1 (9.79) | 59.9 (12.2) | 61.7 (14.9) | 63.4 (11.6) | 63.7 (9.68) | 62.4 (12.5) | 63.2 (13.4) | 61.5 (10.1) | 67.2 (10.4) | 62.9 (11.1) | 64.9 (9.75) | 68.9 (8.31) | 61.2 (11.6) | 64.9 (9.51) | 68.3 (9.91) |
| Median (Q1, Q3) | 64.0 (56.0, 72.0) | 59.0 (49.0, 67.0) | 66.0 (58.0, 72.0) | 60.0 (52.0, 69.0) | 64.0 (53.5, 72.0) | 65.0 (56.0, 71.0) | 64.0 (57.0, 70.0) | 63.0 (57.0, 71.0) | 65.0 (55.0, 73.0) | 62.0 (55.0, 69.0) | 68.0 (60.0, 75.0) | 64.0 (56.0, 71.0) | 66.0 (58.3, 72.0) | 69.0 (63.0, 75.0) | 62.0 (55.0, 70.0) | 65.0 (58.0, 71.0) | 69.0 (62.0, 76.0) |
| **Index Year (Year at First Line of Therapy)** |  |  |  |  |  |  |  |  |  |  |  |  |  |  |  |  |  |
| Mean (SD) | 2016 (2.86) | 2016 (2.47) | 2016 (2.26) | 2015 (3.38) | 2015 (3.38) | 2017 (1.95) | 2017 (1.92) | 2016 (2.30) | 2017 (1.94) | 2014 (3.16) | 2017 (2.09) | 2016 (2.52) | 2017 (1.86) | 2017 (2.04) | 2016 (2.51) | 2016 (2.02) | 2017 (2.04) |
| Median (Q1, Q3) | 2017 (2015, 2018) | 2016 (2014, 2018) | 2016 (2014, 2018) | 2017 (2015, 2018) | 2016 (2014, 2017) | 2017 (2016, 2018) | 2017 (2016, 2018) | 2016 (2015, 2018) | 2017 (2016, 2019) | 2015 (2012, 2017) | 2017 (2016, 2018) | 2016 (2014, 2018) | 2017 (2016, 2019) | 2017 (2015, 2018) | 2016 (2014, 2018) | 2017 (2015, 2018) | 2017 (2015, 2019) |
| **Time from diagnosis to frontline therapy (days)** |  |  |  |  |  |  |  |  |  |  |  |  |  |  |  |  |  |
| Mean (SD) | 740 (1340) | 1690 (2000) | 1910 (2100) | 551 (836) | 114 (718) | 214 (417) | 660 (770) | 357 (598) | 1220 (1720) | 865 (1560) | 318 (644) | 658 (1340) | 262 (465) | 1890 (1940) | 904 (1480) | 81.4 (274) | 634 (1050) |
| Median (Q1, Q3) | 157 (33.0, 836) | 999 (257, 2390) | 1290 (311, 2640) | 161 (38.0, 778) | 18.0 (8.00, 33.0) | 42.0 (25.0, 233) | 447 (69.0, 897) | 140 (41.0, 421) | 611 (86.0, 1530) | 118 (33.0, 872) | 54.0 (29.0, 327) | 49.0 (25.0, 789) | 65.0 (20.0, 367) | 1140 (447, 2830) | 265 (56.0, 1060) | 20.0 (12.0, 32.0) | 261 (61.5, 674) |
| **Gender** |  |  |  |  |  |  |  |  |  |  |  |  |  |  |  |  |  |
| F | 15548 (55.4%) | 4750 (98.9%) | 34 (31.2%) | 2307 (45.6%) | 62 (38.0%) | 423 (24.7%) | 70 (18.2%) | 46 (25.1%) | 237 (33.1%) | 135 (41.5%) | 3601 (50.3%) | 2338 (100%) | 878 (45.8%) | 0 (0%) | 181 (27.9%) | 220 (49.3%) | 266 (30.1%) |
| M | 12531 (44.6%) | 51 (1.1%) | 75 (68.8%) | 2752 (54.4%) | 101 (62.0%) | 1289 (75.3%) | 315 (81.8%) | 137 (74.9%) | 480 (66.9%) | 190 (58.5%) | 3556 (49.7%) | 0 (0%) | 1040 (54.2%) | 1235 (100%) | 467 (72.1%) | 226 (50.7%) | 617 (69.9%) |
| **Race** |  |  |  |  |  |  |  |  |  |  |  |  |  |  |  |  |  |
| White | 20532 (73.1%) | 3407 (71.0%) | 67 (61.5%) | 3579 (70.7%) | 119 (73.0%) | 1235 (72.1%) | 302 (78.4%) | 107 (58.5%) | 613 (85.5%) | 244 (75.1%) | 5215 (72.9%) | 1765 (75.5%) | 1447 (75.4%) | 896 (72.6%) | 498 (76.9%) | 358 (80.3%) | 680 (77.0%) |
| Nonwhite | 7547 (26.9%) | 1394 (29.0%) | 42 (38.5%) | 1480 (29.3%) | 44 (27.0%) | 477 (27.9%) | 83 (21.6%) | 76 (41.5%) | 104 (14.5%) | 81 (24.9%) | 1942 (27.1%) | 573 (24.5%) | 471 (24.6%) | 339 (27.4%) | 150 (23.1%) | 88 (19.7%) | 203 (23.0%) |
| **BMI** |  |  |  |  |  |  |  |  |  |  |  |  |  |  |  |  |  |
| Normal and underweight | 11286 (40.2%) | 1722 (35.9%) | 34 (31.2%) | 1793 (35.4%) | 56 (34.4%) | 817 (47.7%) | 198 (51.4%) | 60 (32.8%) | 195 (27.2%) | 114 (35.1%) | 3288 (45.9%) | 1090 (46.6%) | 956 (49.8%) | 268 (21.7%) | 202 (31.2%) | 163 (36.5%) | 330 (37.4%) |
| Overweight | 9330 (33.2%) | 1514 (31.5%) | 44 (40.4%) | 1679 (33.2%) | 62 (38.0%) | 547 (32.0%) | 127 (33.0%) | 61 (33.3%) | 293 (40.9%) | 115 (35.4%) | 2353 (32.9%) | 661 (28.3%) | 636 (33.2%) | 502 (40.6%) | 240 (37.0%) | 166 (37.2%) | 330 (37.4%) |
| Obese | 7463 (26.6%) | 1565 (32.6%) | 31 (28.4%) | 1587 (31.4%) | 45 (27.6%) | 348 (20.3%) | 60 (15.6%) | 62 (33.9%) | 229 (31.9%) | 96 (29.5%) | 1516 (21.2%) | 587 (25.1%) | 326 (17.0%) | 465 (37.7%) | 206 (31.8%) | 117 (26.2%) | 223 (25.3%) |
| **Smoking status** |  |  |  |  |  |  |  |  |  |  |  |  |  |  |  |  |  |
| History of smoking | 16313 (58.1%) | 1919 (40.0%) | 51 (46.8%) | 2449 (48.4%) | 62 (38.0%) | 1127 (65.8%) | 274 (71.2%) | 125 (68.3%) | 358 (49.9%) | 126 (38.8%) | 5842 (81.6%) | 969 (41.4%) | 991 (51.7%) | 652 (52.8%) | 328 (50.6%) | 429 (96.2%) | 611 (69.2%) |
| No history of smoking | 11766 (41.9%) | 2882 (60.0%) | 58 (53.2%) | 2610 (51.6%) | 101 (62.0%) | 585 (34.2%) | 111 (28.8%) | 58 (31.7%) | 359 (50.1%) | 199 (61.2%) | 1315 (18.4%) | 1369 (58.6%) | 927 (48.3%) | 583 (47.2%) | 320 (49.4%) | 17 (3.8%) | 272 (30.8%) |
| **TP53 (Short Variant) status** |  |  |  |  |  |  |  |  |  |  |  |  |  |  |  |  |  |
| 0 | 10521 (37.5%) | 2452 (51.1%) | 73 (67.0%) | 1216 (24.0%) | 97 (59.5%) | 401 (23.4%) | 198 (51.4%) | 118 (64.5%) | 533 (74.3%) | 278 (85.5%) | 2571 (35.9%) | 450 (19.2%) | 488 (25.4%) | 688 (55.7%) | 552 (85.2%) | 36 (8.1%) | 370 (41.9%) |
| 1 | 17558 (62.5%) | 2349 (48.9%) | 36 (33.0%) | 3843 (76.0%) | 66 (40.5%) | 1311 (76.6%) | 187 (48.6%) | 65 (35.5%) | 184 (25.7%) | 47 (14.5%) | 4586 (64.1%) | 1888 (80.8%) | 1430 (74.6%) | 547 (44.3%) | 96 (14.8%) | 410 (91.9%) | 513 (58.1%) |
| **Tissue TMB score** |  |  |  |  |  |  |  |  |  |  |  |  |  |  |  |  |  |
| Mean (SD) | 6.98 (13.7) | 4.85 (8.53) | 2.31 (4.47) | 6.04 (13.3) | 11.9 (10.2) | 6.13 (7.94) | 6.10 (8.41) | 4.72 (9.48) | 27.9 (36.5) | 4.96 (7.70) | 9.47 (15.0) | 3.75 (11.8) | 4.04 (7.69) | 4.92 (11.4) | 3.44 (3.71) | 8.83 (6.86) | 9.17 (13.8) |
| Median (Q1, Q3) | 3.75 (1.74, 7.50) | 2.61 (1.25, 5.22) | 1.61 (0.807, 2.42) | 3.48 (2.40, 5.22) | 8.88 (4.84, 15.3) | 4.35 (2.50, 6.96) | 3.75 (2.40, 7.50) | 3.48 (1.74, 5.22) | 15.6 (5.22, 34.8) | 3.23 (1.25, 6.46) | 6.25 (2.61, 11.3) | 2.61 (1.25, 4.35) | 2.50 (1.25, 3.75) | 2.50 (1.25, 4.35) | 2.61 (1.25, 4.35) | 7.50 (4.85, 11.3) | 6.25 (3.48, 11.3) |
| **PDL1 status** |  |  |  |  |  |  |  |  |  |  |  |  |  |  |  |  |  |
| 0 | 25913 (92.3%) | 4639 (96.6%) | 99 (90.8%) | 4690 (92.7%) | 148 (90.8%) | 1395 (81.5%) | 322 (83.6%) | 168 (91.8%) | 644 (89.8%) | 297 (91.4%) | 6673 (93.2%) | 2240 (95.8%) | 1789 (93.3%) | 1112 (90.0%) | 601 (92.7%) | 418 (93.7%) | 678 (76.8%) |
| 1 | 2166 (7.7%) | 162 (3.4%) | 10 (9.2%) | 369 (7.3%) | 15 (9.2%) | 317 (18.5%) | 63 (16.4%) | 15 (8.2%) | 73 (10.2%) | 28 (8.6%) | 484 (6.8%) | 98 (4.2%) | 129 (6.7%) | 123 (10.0%) | 47 (7.3%) | 28 (6.3%) | 205 (23.2%) |
| **Last Albumin, Abnormal** |  |  |  |  |  |  |  |  |  |  |  |  |  |  |  |  |  |
| no | 24245 (86.3%) | 4479 (93.3%) | 104 (95.4%) | 4392 (86.8%) | 144 (88.3%) | 1379 (80.5%) | 335 (87.0%) | 134 (73.2%) | 649 (90.5%) | 280 (86.2%) | 5928 (82.8%) | 1985 (84.9%) | 1539 (80.2%) | 1179 (95.5%) | 567 (87.5%) | 389 (87.2%) | 762 (86.3%) |
| yes | 3834 (13.7%) | 322 (6.7%) | 5 (4.6%) | 667 (13.2%) | 19 (11.7%) | 333 (19.5%) | 50 (13.0%) | 49 (26.8%) | 68 (9.5%) | 45 (13.8%) | 1229 (17.2%) | 353 (15.1%) | 379 (19.8%) | 56 (4.5%) | 81 (12.5%) | 57 (12.8%) | 121 (13.7%) |
| **Last Absolute Lymphocyte Count, Abnormal** |  |  |  |  |  |  |  |  |  |  |  |  |  |  |  |  |  |
| no | 20421 (72.7%) | 3645 (75.9%) | 37 (33.9%) | 3969 (78.5%) | 103 (63.2%) | 1186 (69.3%) | 209 (54.3%) | 130 (71.0%) | 552 (77.0%) | 214 (65.8%) | 4827 (67.4%) | 1752 (74.9%) | 1405 (73.3%) | 893 (72.3%) | 498 (76.9%) | 363 (81.4%) | 638 (72.3%) |
| yes | 7658 (27.3%) | 1156 (24.1%) | 72 (66.1%) | 1090 (21.5%) | 60 (36.8%) | 526 (30.7%) | 176 (45.7%) | 53 (29.0%) | 165 (23.0%) | 111 (34.2%) | 2330 (32.6%) | 586 (25.1%) | 513 (26.7%) | 342 (27.7%) | 150 (23.1%) | 83 (18.6%) | 245 (27.7%) |

Given patient heterogeneity and vast differences in sample sizes of the different cancer cohorts, we sought to investigate whether models developed on the pan-cancer cohort could improve survival predictions compared to those developed in single-cancer settings, by learning from all of the available information and potential signals across cancer types. Using the full high-dimensional, clinico-genomic feature set, we developed a series of penalized Cox proportional hazards models (“Full” models) to predict survival from 1L initiation date in (i) the large pan-cancer cohort (*pan-cancer* model) and (ii) each of the 16 cancer cohorts separately (*single-cancer* models). To compare to single-cancer models, pan-cancer trained models were evaluated on each of the 16 separate cancer cohorts in addition to the pan-cancer cohort (Figure 1). We benchmarked these high-dimensional prognostic models against simpler models from clinical practice and the literature, and here we present: (1) a “benchmark” model containing cancer type, age, gender, race, smoking status, cancer stage at diagnosis, baseline Eastern Cooperative Oncology Group (ECOG) Performance Status, time from diagnosis to initiation of 1L, and time from genomic test to initiation of 1L; and (2) a model adapted from ROPRO (Real wOrld PROgnostic score) by Becker *et al^9^*, referred to as “ROPRO-like”. These models are described in Table S2 and SI Materials & Methods.

The out-of-sample performance of each trained pan-cancer and single-cancer prognostic model was evaluated on a withheld, single-cancer test dataset containing 20% of the total patient cohort (split by stratified random sampling on cancer type), using three performance metrics to assess the discrimination and calibration of the survival predictions: concordance index (c-index), integrated Brier score (IBS), and the hazard ratio (HR) comparing survival between patients in predicted low-risk and high-risk groups based upon a median split. Bias-corrected 95% confidence intervals for the c-index and IBS were obtained via 1,000 bootstrap replicates of the train and test data^16^.

### Model performance in pan- and single-cancer cohorts

Table 2 summarizes the out-of-sample c- index for the different comparator models evaluated on each cancer cohort. The c-index provides a measure of how well a model can discriminate prognosis between patients. For each model, the pan-cancer out-of-sample performance is presented alongside that of the equivalent single-cancer model. Figure 2 visualizes trends in pan-cancer and single-cancer c-indexes across (A) all 3 comparator models and (B) all 16 cancer cohorts for only the full high-dimensional model.

**Figure 2.**
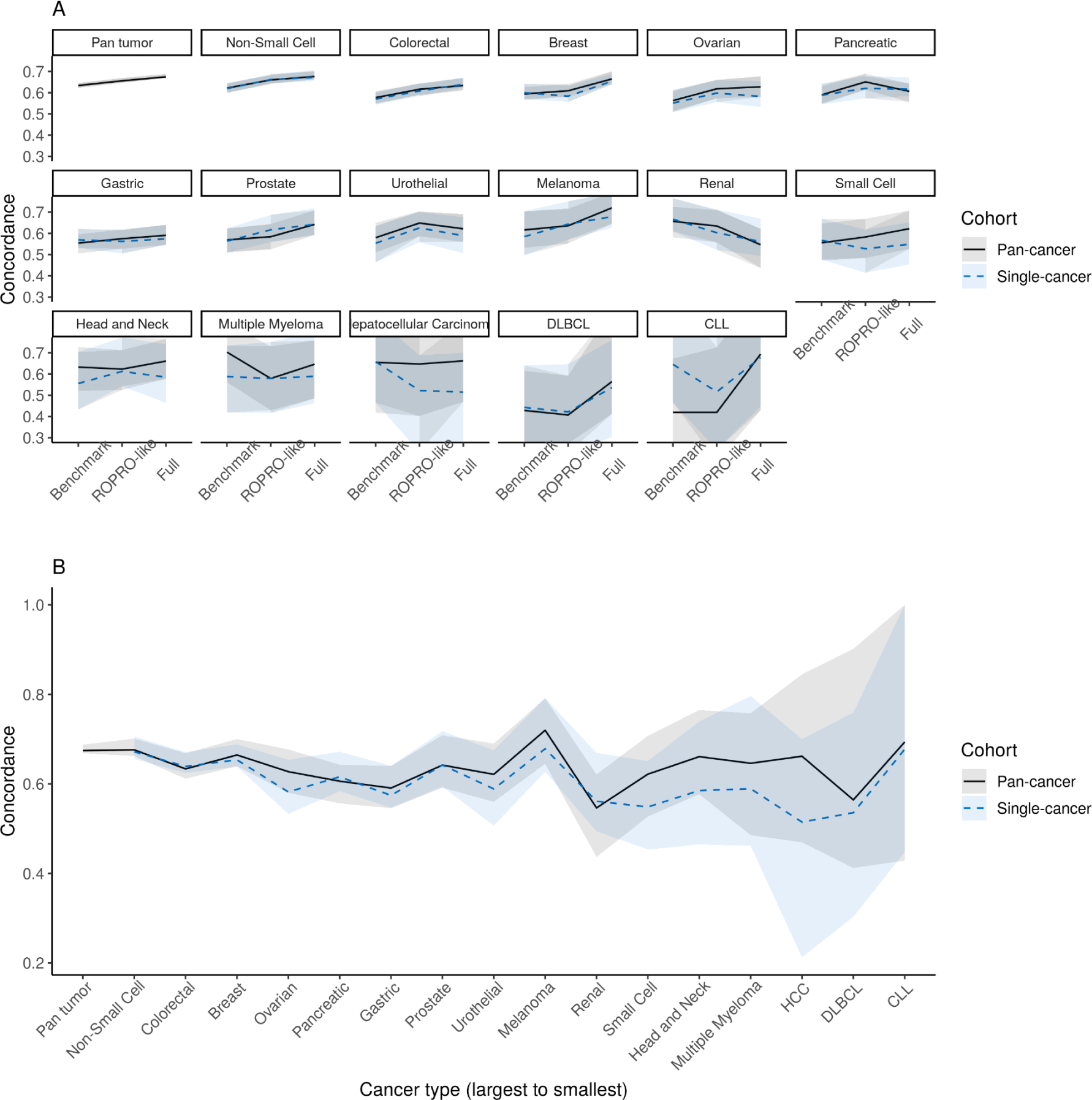
The out-of-sample concordance index (c-index), where values closer to 1 indicate higher prognostic discrimination. (A) The pan-cancer (solid black line) and single-cancer (dashed blue line) c-index for each comparator model of increasing high-dimensionality (x-axis: Benchmark, ROPRO-like, and Full models). (B) For each cancer cohort, we compare the pan-cancer (solid black line) and single-cancer (dashed blue line) c-index for the Full model constructed on the full feature set of > 2,000 features. 95% bias-corrected percentile intervals (shaded) around the estimates are shown for 1,000 bootstrap replicates. In both plots, cancer types are arranged from largest to smallest sample size.

**Table 2.** C-index performance measures for single-cancer (SC) and pan-cancer (PC) models of increasing number of predictors (‘p’).

| Indication | Number of events (%) | Comparator models |  |  |  |  |  |
| --- | --- | --- | --- | --- | --- | --- | --- |
|  |  | Benchmark (p = 9) |  | RoPro-like (p = 29) |  | Full Model (p = 2,109) |  |
|  |  | SC | PC | SC | PC | SC | PC |
| Pan cancer (n = 28,079) |  | N/A | 0.63 (0.62,0.65) | N/A | 0.66 (0.65,0.67) | N/A | 0.673 (0.667,0.687) |
| Non-Small Cell Lung Cancer (n = 7,157) | 4,137 (57.8%) | 0.62 (0.6,0.64) | 0.62 (0.6,0.65) | 0.66 (0.64,0.69) | 0.66 (0.64,0.68) | 0.67 (0.66,0.7) | 0.67 (0.66,0.7) |
| Colorectal (n = 5,059) | 2,751 (54.4%) | 0.57 (0.54,0.6) | 0.58 (0.55,0.61) | 0.61 (0.58,0.64) | 0.62 (0.59,0.64) | 0.64 (0.62,0.68) | 0.63 (0.61,0.67) |
| Breast (n = 4,801) | 2,412 (50.2%) | 0.6 (0.57,0.64) | 0.59 (0.56,0.63) | 0.58 (0.56,0.63) | 0.61 (0.58,0.64) | 0.66 (0.64,0.7) | 0.66 (0.64,0.7) |
| Ovarian (n = 2,338) | 1,063 (45.5%) | 0.55 (0.51,0.61) | 0.56 (0.51,0.61) | 0.6 (0.56,0.66) | 0.62 (0.57,0.66) | 0.57 (0.52,0.63) | 0.62 (0.57,0.67) |
| Pancreatic (n = 1,918) | 1,349 (70.3%) | 0.59 (0.54,0.64) | 0.59 (0.55,0.63) | 0.62 (0.57,0.68) | 0.65 (0.61,0.69) | 0.62 (0.58,0.67) | 0.61 (0.57,0.65) |
| Gastric (n = 1,712) | 1,131 (66.1%) | 0.57 (0.53,0.62) | 0.55 (0.51,0.6) | 0.56 (0.51,0.62) | 0.57 (0.53,0.62) | 0.57 (0.54,0.63) | 0.58 (0.54,0.63) |
| Prostate (n = 1,235) | 633 (51.3%) | 0.56 (0.51,0.62) | 0.57 (0.51,0.62) | 0.62 (0.56,0.69) | 0.58 (0.52,0.64) | 0.65 (0.62,0.72) | 0.63 (0.57,0.69) |
| Urothelial (n = 883) | 527 (59.7%) | 0.55 (0.46,0.63) | 0.58 (0.51,0.65) | 0.63 (0.56,0.7) | 0.65 (0.59,0.7) | 0.6 (0.52,0.67) | 0.62 (0.55,0.68) |
| Melanoma (n = 717) | 315 (43.9%) | 0.58 (0.5,0.7) | 0.62 (0.53,0.7) | 0.64 (0.57,0.75) | 0.64 (0.55,0.72) | 0.66 (0.58,0.77) | 0.72 (0.65,0.8) |
| Renal (n = 648) | 332 (51.2%) | 0.67 (0.61,0.77) | 0.66 (0.58,0.72) | 0.6 (0.52,0.71) | 0.63 (0.56,0.71) | 0.48 (0.36,0.57) | 0.55 (0.44,0.64) |
| Small Cell Lung Cancer (n = 446) | 321 (72.0%) | 0.57 (0.48,0.67) | 0.56 (0.47,0.65) | 0.53 (0.41,0.62) | 0.58 (0.48,0.67) | 0.51 (0.39,0.59) | 0.63 (0.53,0.71) |
| Head and Neck (n = 385) | 237 (61.6%) | 0.56 (0.43,0.7) | 0.63 (0.52,0.73) | 0.61 (0.54,0.77) | 0.62 (0.53,0.71) | 0.58 (0.45,0.73) | 0.66 (0.57,0.76) |
| Multiple Myeloma (n = 325) | 138 (42.5%) | 0.59 (0.42,0.73) | 0.7 (0.56,0.84) | 0.58 (0.42,0.75) | 0.58 (0.43,0.73) | 0.58 (0.43,0.77) | 0.63 (0.44,0.75) |
| Hepatocellular Carcinoma (n = 183) | 112 (61.2%) | 0.66 (0.47,0.86) | 0.65 (0.42,0.84) | 0.52 (0.23,0.69) | 0.65 (0.4,0.81) | 0.6 (0.32,0.82) | 0.59 (0.36,0.75) |
| DLBCL (n = 163) | 70 (42.9%) | 0.44 (0.22,0.64) | 0.43 (0.25,0.62) | 0.42 (0.23,0.65) | 0.41 (0.27,0.59) | 0.54 (0.32,0.79) | 0.57 (0.43,0.88) |
| CLL (n = 109) | 24 (22.0%) | 0.65 (0.47,1) | 0.42 (0.16,0.67) | 0.52 (0.24,0.86) | 0.42 (0.19,0.72) | 0.69 (0.5,1) | 0.66 (0.36,1) |

*Large vs. small feature set performance:* Across comparator models (benchmark, ROPRO-like, and full), there was a consistent gain in discrimination (higher c-index) for most cancer types as additional features were incorporated into the model, although this had diminishing returns for cancer cohorts with fewer patients (Fig. 2A). The overall pan-cancer c-index improved slightly from the benchmark model (0.63, 95% CI: 0.62, 0.65) to the full model (0.673, 95% CI: 0.667, 0.687) with even more modest gains over the ROPRO-like model (0.66, 95% CI: 0.65, 0.67) (Table 2). Single-cancer models followed similar trends. The change in c-index was small relative to the uncertainty resulting from small sample sizes in the test set, so these findings are descriptive. However, some of the smallest cancer cohorts - with fewer than 500 patients in the training data, e.g. small-cell lung cancer, multiple myeloma, and hepatocellular carcinoma (HCC) - saw little or no improvement in the c-index as the number of features increased. These findings suggest a possible performance trade-off between sample size and number of predictors.

*Pan-vs. single-cancer performance:* In almost all cases, however, the pan-cancer model performed similarly to or outperformed the equivalent single-cancer models (trained on the same predictors) on the basis of the c-index, most substantially in the full (highest-dimensional) model (Fig. 2B, Table 2). In particular for the full model, c-index improvements were highest among the smallest sample sizes, ranging from 2-20% higher in cancer cohorts with less than 500 patients. Meaningful gains were not observed for cancer cohorts with large sample sizes. It is worth noting that the uncertainty around these estimates is considerably high as a result of very small sample sizes, so findings are descriptive and this is observed as a general trend.

To further assess the prognostic discrimination of these models, we calculated a prognostic score for each patient using the final coefficients of each penalized Cox model, representing the predicted risk of death. We then stratified patients by cancer type into low- and high-risk categories based on the median score of the training patients in each cancer cohort.

Aligning with trends in the c-index and sample size, the pan-cancer model outperformed single-cancer models on patient risk stratification for many cancers, most prominently for cancers with smaller sample sizes. Fig. 3A visualizes key factors driving risk stratification in the pan-cancer model, such as lab tests, year of frontline therapy, tissue tumor mutational burden (tTMB), ECOG score, and TP53 (SV) alteration. In Fig. 3B, the pan-cancer model yielded clear separation of the survival curve between high- and low-risk patients in the pan-cancer cohort, and training on the pan-cancer dataset generally yielded an improvement over single-cancer models (Fig. 3C-F, Figs. S1-5). For 12 of the 16 cancer types, hazard ratios (HRs) comparing the survival of high to low-risk patients were lower, many with narrower confidence intervals and more well-separated survival curves, using pan-cancer predictions compared to cancer-specific predictions; for example in ovarian cancer (OC), the HR was 0.48 (95% CI: 0.34, 0.66) in the pan-cancer model (Fig. 3C) compared to 0.74 (95% CI: 0.55, 0.99) in the single-cancer model (Fig. 3D). In small-cell lung cancer (SCLC), the pan- and single-cancer HRs were 0.48 (95% CI: 0.29, 0.78) versus 1 (95% CI: 0.62, 1.62), respectively (Fig. 3E-F).

**Figure 3.**
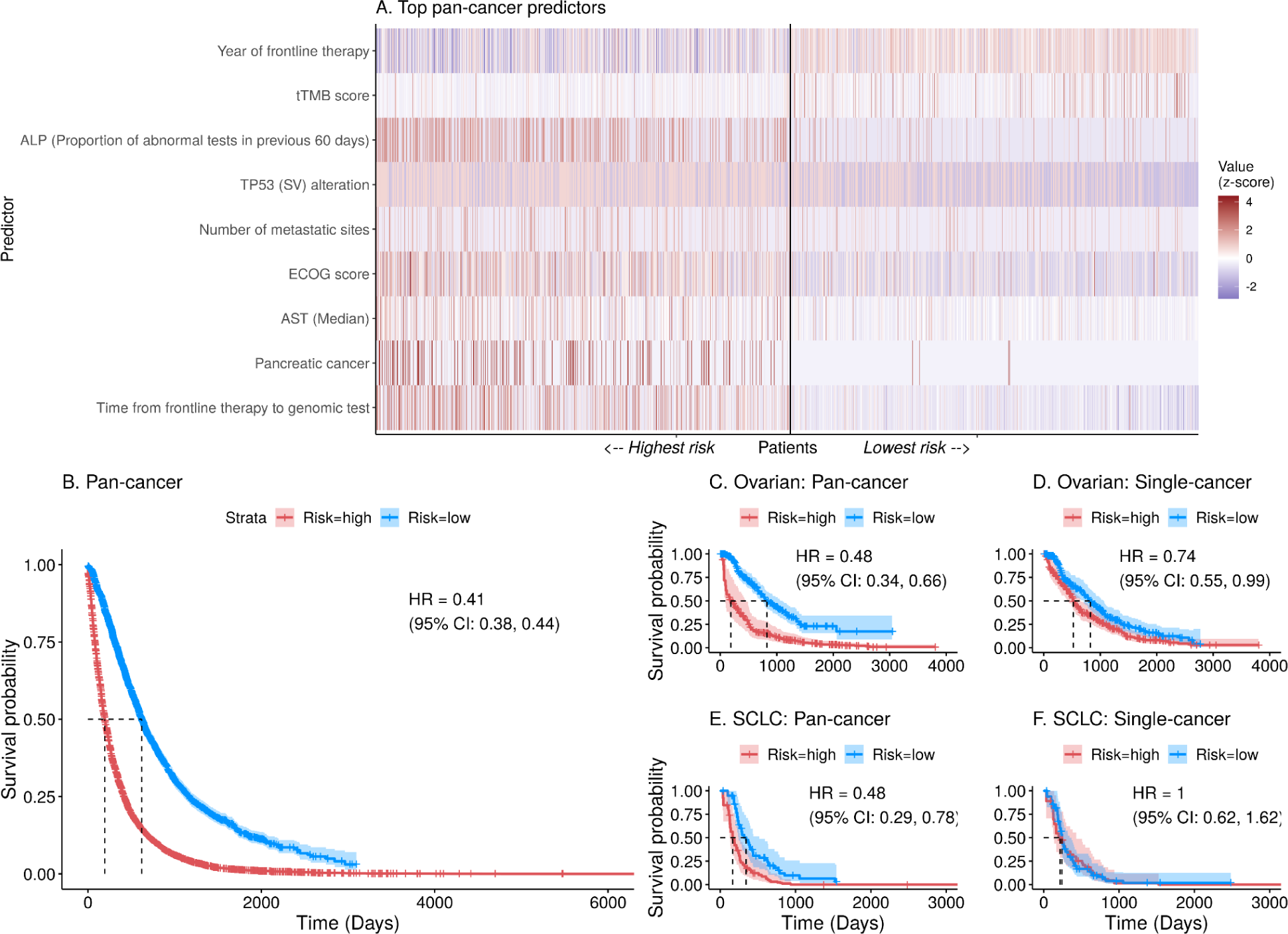
(A) Heatmap showing the normalized (z-score) value of selected pan-cancer predictors in the highest (top 25%) and lowest (bottom 25%) risk patients in the out-of-sample pan-cancer cohort. (B) Out-of-sample pan-cancer risk stratification. In single-cancer cohorts, risk stratification of pan- and single-cancer models are shown for (C-D) Ovarian Cancer and (E-F) Small Cell Lung Cancer (SCLC). In (A), from left to right, patients are ordered from highest to lowest risk based on their risk scores, and their normalized value for each predictor is given in shades of red (higher) or blue (lower). A vertical line separates highest from lowest risk patients.

Trends in the IBS, a measure of performance reflecting both discrimination and calibration, were less clear and presented in Fig. S6. Taken together with the c-index results, the IBS results suggest that single-cancer predictions tended to be slightly better calibrated than pan-cancer predictions, in particular for the largest cancer cohorts. Similar to the c-index, the IBS improved with additional features, where improvement is marked by decreases in the score; however, the range of possible values of IBS was smaller than the uncertainty around each estimate, making interpretation difficult.

### Clinico-genomic factors associated with cancer survival

Our high-dimensional pan- and single-cancer models offer the opportunity to assess variables strongly associated with cancer prognosis, out of all variables available in the clinico-genomic database. Our penalized approach using lasso regularization provides feature selection by shrinking the coefficients of unimportant variables to zero and retaining only the most prognostic features in the model. Of all 2,059 predictors (2,135 after one-hot encoding), the pan-cancer full model selected a total of 354. Figure 4 shows the coefficients of the top 25 pan-cancer predictors, interpreted as the log hazard ratio (HR) where positive coefficients indicate worse prognosis (harmful association with survival) and negative coefficients indicate better prognosis (favorable association with survival). Several of these predictors were described in the previous section as having contributed to the effective pan-cancer risk stratification (Fig. 3A). Notably, the clinical features associated with substantially worse survival (log HR > 0.09) were longer time from frontline therapy to genomic test, pancreatic and gastric cancer types, higher ECOG score, higher aspartate aminotransferase (AST) levels, and higher heart rate. Clinical features associated with substantially better survival (log HR < -0.09) were more recent year of frontline treatment and higher albumin levels. Indicators for cancer type suggested pronounced differences in cancer-specific survival: gastric and pancreatic were strongly associated with worse survival, whereas CLL, a slow-growing blood cancer, was associated with longer survival. Seven genomic mutations were among the top 25 pan-cancer predictors overall: KDM6A (CN), AR (CN), KEAP1 (SV), PAX5 (RE), and TP53 (SV) (all associated with worse survival), and FGFR4 (SV) and ALK (RE) (both associated with better survival). In addition, higher tissue tumor mutational burden (tTMB) was associated with better survival.

**Figure 4.**
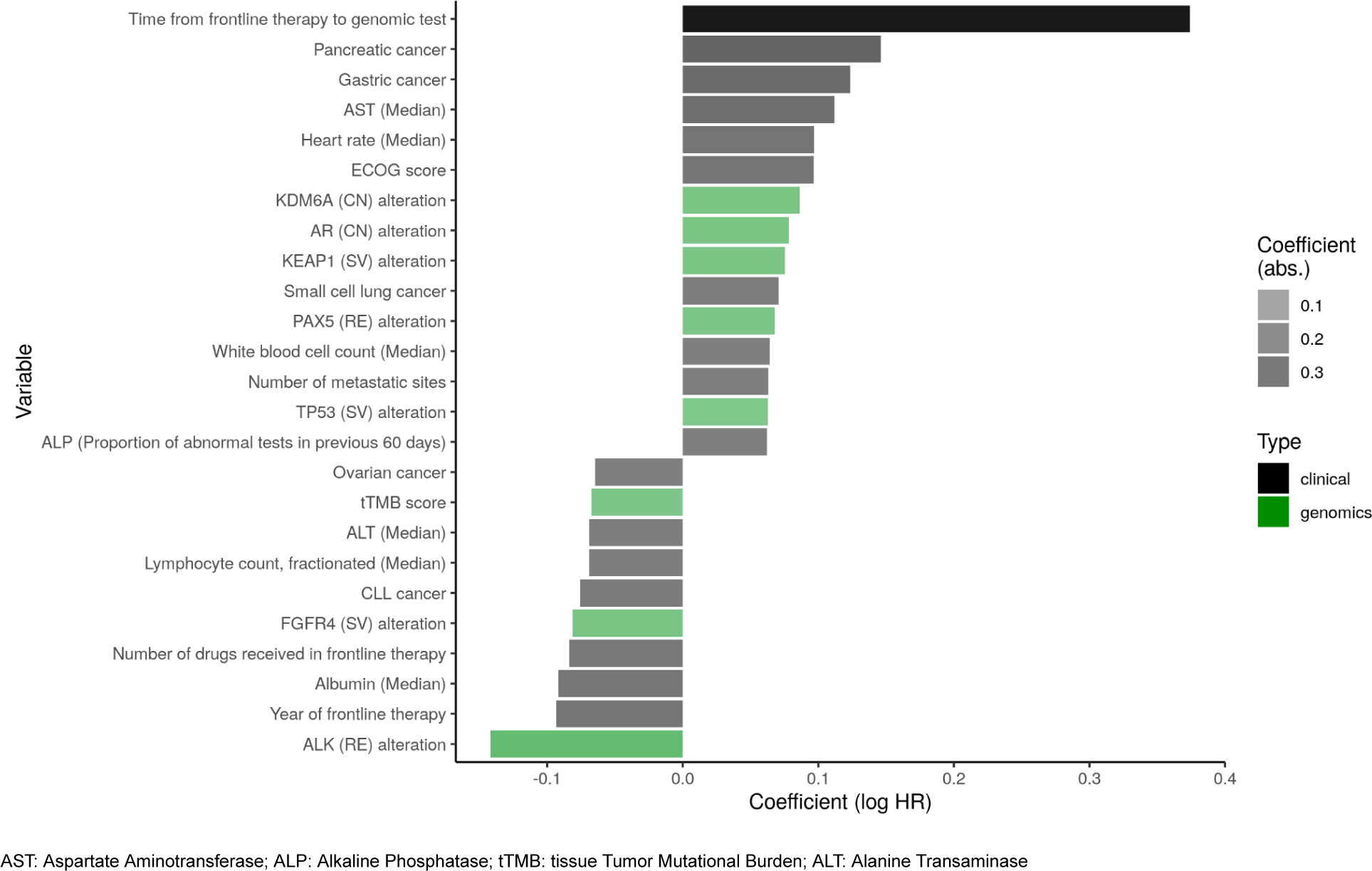
The top 25 predictors selected by the full penalized pan-cancer model, ordered and shaded by coefficient size (log hazard ratio, HR) and colored by predictor type (black = clinical, green = genomics). Positive coefficients suggest a more harmful association with survival and negative coefficients suggest a more favorable association.

Our single-cancer models, trained on the equivalent feature set of over 2,000 clinico-genomic predictors but learning from only patients of a single cancer type, offered insights into features important in each cancer setting independently. In contrast with the pan-cancer model, genomic features were more commonly selected as top predictors in the single-cancer models and revealed unique cancer-specific genomic profiles with mutations or cancer pathways not identified as top predictors in the pan-cancer model. Figs. S7-9 show these top predictors for each of the 16 cancer types assessed.

In addition, evaluating the clinical and genomic predictors selected by multiple single-cancer models could reveal relationships between cancer types and corroborate findings of the pan-cancer model. Fig. 5 shows the coefficients of (A) the top 25 pan-cancer variables and (B) the 15 clinico-genomic variables that were selected by at least 7 pan- and single-cancer models. Strong consistency in effect size and direction was observed across several cancers, including for the genomic variables presence of a TP53 (SV) mutation and higher tumor purity (both associated with worse survival); and for the clinical variables older age, higher ECOG score, higher heart rate, and higher proportion of abnormal results for lab tests like alkaline phosphatase (ALP), albumin, and lymphocyte count (all associated with worse survival). Across most cancer cohorts, there was also consistency in the effects of 2 temporal variables: year of frontline therapy (more recent years are associated with better survival) and time from frontline treatment to genomic test (longer interval is associated with worse survival).

**Figure 5.**
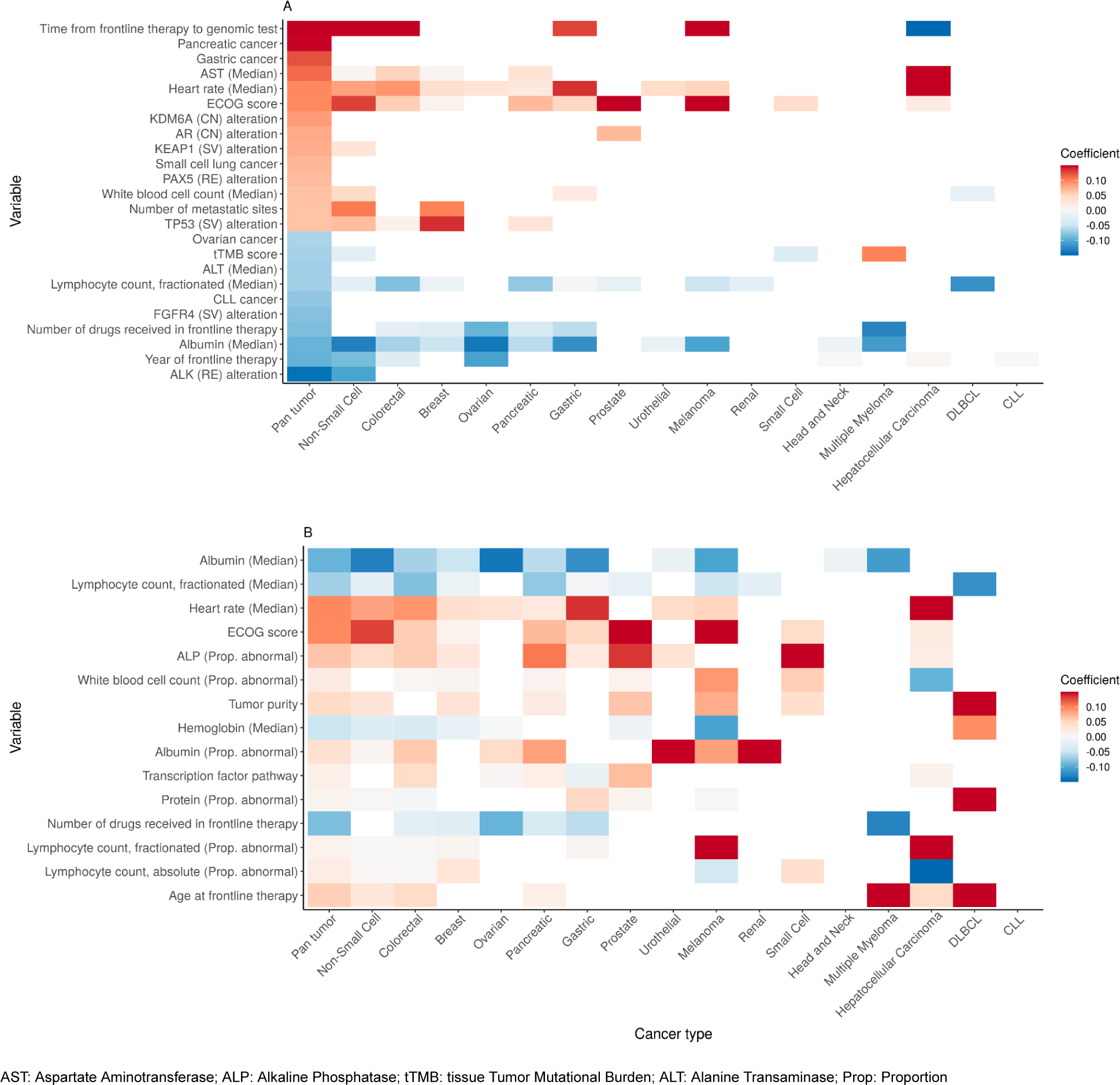
Heatmaps showing the coefficients, by cancer setting, of (A) the top 25 pan-cancer variables and (B) the 15 variables that were selected by at least 7 single-cancer models. In (A), variables on the y-axis are arranged by descending coefficient in the pan-cancer model (high to low); in (B), by descending frequency of selection in the single-cancer models (most to fewest). Variables that were not selected in a particular cancer setting are represented as blank (white) tiles. Cancer types are arranged on the x-axis from largest to smallest sample size.

Most interesting, perhaps, are the predictors selected exclusively by the pan-cancer model and which appear in the model’s top 25 variables (Fig. 5A): the mutations in KDM6A (CN), PAX5 (RE), and FGFR4 (SV). These mutations all exhibited stronger association with survival compared to TP53 (SV, the most frequent mutation across all patients), with log HRs between 0.07-0.14 in absolute value, yet were not selected by any cancer-specific model.

Finally, we note that the sparsity of the single-cancer models — how many variables were selected in each cancer cohort — is related to their sample size. Because variables are shrunken by the model to prevent overfitting, the smallest cancer subgroups are often constrained from selecting as many variables as the larger subgroups (Fig. S10). As a result, the present feature analysis considers a limited interpretation of variable selection: variables selected by models are considered informative, but the absence of variables is not necessarily informative and may be instead a consequence of sample size.

## Discussion

Our systematic analysis of prognostic pan-cancer and single-cancer models demonstrates the value of a comprehensive framework for prognostic modeling, namely around: model performance tradeoffs, shared cancer biologies, and novel pan-cancer predictors.

First, we observed that, compared to single-cancer models, pan-cancer models demonstrated improved performance and risk stratification capabilities in many cancer types, specifically when the sample size and event rate was small and the training set had a large number of predictors. In the highest-dimensional models (the “full” model), the ability to train on an extensive number of clinico-genomic factors across multiple cancer types was a learning advantage of the pan-cancer model, allowing it to select a vast number of predictors compared to the equivalent single-cancer model, which could be considered an example of transfer learning in low-data settings^17^. Transfer learning assumes that predictive features learned from training in some domain can be applied to a different domain - in this case, cancer types. This is apparent when studying the performance differences between the pan-cancer and single-cancer models. For the largest cancer cohorts (n > 1,000 patients) like NSCLC and breast cancer, little difference was made because both the pan-cancer and single-cancer datasets were sufficiently large to train on many relevant predictors. For example, in NSCLC (n > 5,000 patients), the single-cancer model selected 115 variables compared to the pan-cancer model’s selection of 354. Moreover, the large cancer single-cancer models shared many of the same predictors selected by the pan-cancer model, such as metastatic sites, ALK mutation (specific to NSCLC), and estrogen receptor (ER) positive status (specific to breast cancer). However, for the smallest cancer cohorts, the models were penalized resulting in the tendency to select fewer features. For head and neck cancer (n = 319 patients), the single-cancer model selected only 6 variables out of > 2,000, but its performance markedly improved with the pan-cancer model which ultimately selected 354 variables. This learning advantage of the pan-cancer model allowed it to capture a wide range of prognostic factors to apply to prediction and risk stratification, especially in smaller cancer settings.

However, pan-cancer models did not show a learning advantage in simpler, lower-dimensional settings. In our comparator models (benchmark and ROPRO-like models) which contained a considerably smaller number (< 30) of features, performance was more similar between pan- and single-cancer models. In these scenarios, data from multiple cancer types may not provide substantial additional information or predictive power over what is available in the single cancer setting, particularly since these models already include a small number of known highly prognostic factors such as age, ECOG score, and lab tests. Further, these simple models still perform quite well thanks to the inclusion of ECOG, which was identified by the full (highest-dimensional) model as a top 10 predictor out of > 2,000 features: even the simplest pan-cancer benchmark model, which included ECOG, achieved a c-index of 0.63 (95% CI: 0.62, 0.65) compared to the full model’s c-index of 0.673 (95% CI: 0.667, 0.687). These results suggest that prognostic models developed using a handful of variables collected in routine clinical practice may be sufficient for certain applications, with the benefit of being easier to implement. Indeed, smaller cancer cohorts did not see much performance improvement moving from simple to complex models; however, larger cancer cohorts with > 500 patients like colorectal, prostate, and melanoma saw marked improvements with the inclusion of additional predictors. Here, the decision to use high-dimensional models should consider the disease setting and weigh tradeoffs including: the impact of the performance increase, the feasibility of collecting more data, the desire to include additional clinico-genomic information, and the need for more computationally intensive processes.

Second, our study revealed strong consistency in the predictors identified by both pan-cancer and single-cancer models, underscoring the presence of common clinical and genomic features that contribute to cancer prognosis and corroborating known disease biology. Across all comparator models and across pan-cancer and single-cancer settings, demographic and clinical features like age, ECOG score, and cancer stage at diagnosis were found to be highly prognostic, with older age, higher ECOG score, higher proportion of abnormal lab results, higher AST, and higher heart rate all associated with worse survival outcomes, echoing what is extensively published in the literature and aligning with the findings of the ROPRO model ^9,18–22^. Other lab tests like higher albumin levels, higher lymphocyte count, and higher hemoglobin (non-anemic status) were consistently associated with better survival, also aligning with literature and the ROPRO model^23–26^. Briefly, low hemoglobin is a marker of anemia, indicating insufficient oxygen transport to tissues, compromising immune response^27^; low albumin reflects poor nutritional status, impairing the body’s ability to fight cancer and recover from treatment^23^; and low lymphocyte count signifies weakened immune surveillance, reducing the body’s capacity to detect and destroy cancer cells^26^. Certain factors related to care were also associated with better survival outcomes across many cancer types including receiving a higher number of drugs in frontline therapy and being treated in more recent years.

Several genomic factors also exhibited strong consistency across cancer types. Higher tumor purity was consistently associated with worse survival in 7 cancer types. The concept of tumor purity refers to the extent to which the tumor tissue consists of cancer cells versus other types of cells present in the tumor microenvironment; a higher tumor purity indicates a larger proportion of cancer cells relative to non-cancerous cells, and here is measured in silico (using epigenomic, genomic, or transcriptomic profiles)^28^. Multiple factors contribute to the tumor purity estimate, including ease of sampling and specific tissue of origin, but an intriguing hypothesis that could explain the effect observed in these data is immune infiltration. The immune system plays a crucial role in recognizing and eliminating abnormal or cancerous cells, but when tumor purity is high, this functionality is inhibited. Another possibility is that samples with higher tumor purity are representative of larger or more aggressive tumors with poor survival. Thus, the finding that high tumor purity is a strong predictor of worse survival is likely aligned with our understanding of cancer biology^29^. Moreover, the TP53 (SV) mutation was associated with worse survival in 5 cancer settings, consistent with the well-established role of TP53 in tumor suppression and its implications in cancer progression^30^.

A new finding was the strong effect of time from frontline therapy to genomic test, across cancers. The left truncation-adjusted models adjusted for this variable to achieve quasi-independence between database entry time (marked by receiving a genomic test) and survival time, described in Materials & Methods. This feature is an important phenomenon in the data, where many patients receive genomic tests long after frontline treatment, and could have implications for patient outcomes. One hypothesis to explain its strong association is that patients who receive genomic tests later in their treatment course may have exhausted standard treatment options. As a result, they may be considered high-risk with limited therapeutic options. This finding raises some considerations for clinical practice: earlier genomic testing in the treatment course may provide more personalized treatment options (e.g. biomarker targeted therapy) for patients, potentially leading to better outcomes. Further, the time interval between diagnosis and genomic test is shrinking over time thanks to the increased availability and access to genomic testing in recent years, so this association may weaken in the future.

Finally, our analysis uncovered potentially unique pan-cancer variables. The somatic mutations in KDM6A (CN), PAX5 (RE), and FGFR4 (SV), identified among the top 25 features of the pan-cancer model, had similar prevalence across multiple cancer types but were not selected by any of the single-cancer models and thus warrant further research. PAX5 may be implicated in metastasis^31^, KDM6A in DNA damage repair, and the FGFR family of proteins in a number of cell proliferation pathways^32^. The predominant rearrangement involving PAX5 was truncation, with a minority of cases being intra-gene rearrangements - no recurrent rearrangements involving non-PAX5 partner genes were observed. Copy number variants of KDM6A were predominantly deletions, except for cases in breast cancer and ovarian cancer, where a range of amplifications was observed. Previously, loss of KDM6A has been associated with poor prognosis in pancreatic cancer ^33^.

While not the focus of this paper, our study also reveals the unique clinico-genomic profile of each cancer type by showcasing the top 25 features of each single-cancer model, which can be explored in Figs. S7-9.

Prognostic models hold significant utility in oncology research and clinical practice. Better risk stratification on the basis of prognosis can help physicians and clinical trials identify high-risk patients who may benefit from more intensive interventions or personalized treatment strategies; conversely, they can also spare low-risk patients from unnecessary interventions and thus help optimize resource allocation. In many cancer settings, we observed that pan-cancer models improved risk stratification because of their training advantages discussed previously. Separately, our models also corroborate the good discriminative ability of the ROPRO prognostic model in both pan- and single-cancer settings, which has demonstrated clinical utility in several applications ^9,34^.

Our prognostic modeling framework is a strength of this work that enables a consistent evaluation of multiple models in diverse pan- and single-cancer settings. It can be used as a template to extend this study to future research areas, including: further work on the theory behind “low information” transfer learning approaches like the one demonstrated in this study; the concept of “pre-training” to give advantage in low information settings; and exploring cancer subgroups (such as hematological or hormone-dependent) within which similarities can be further exploited by this “pan” training approach.

Our study has several limitations that should be considered when interpreting the results. The sample sizes and number of events in many cancer types were relatively small, which can lead to model instability and imprecise estimates, as indicated by the wide confidence intervals for the smallest cancer cohorts like DLBCL (n = 122) and CLL (n = 83) as well as unstable coefficient sizes and very sparse models. As a result, findings that compare model performance remain trends and are descriptive. Additionally, the analysis used penalized linear Cox models, which assume a linear relationship between predictors and the log-hazard ratio. While this is a commonly used approach, it may not capture potential non-linear associations between predictors and outcomes nor complex interactions between variables. To investigate whether the ability to model complex and nonlinear associations and interactions would improve performance, we implemented a tree-based approach in the form of random survival forests adjusted for left truncation^35^ and found no difference in performance compared to our linear models (Fig. S11, SI Materials & Methods), which suggests the data may be adequately modeled using linearity assumptions, which also yield more interpretable results (hazard ratios) for clinical audiences. Our models adjusted for left truncation, a feature of the clinico-genomic database, but this could introduce bias if the truncation is dependent on the outcome. Further, high levels of missingness in the clinico-genomic database led us to omit potential prognostic factors like lactate dehydrogenase (LDH), preventing us from perfectly replicating the ROPRO model^9^ and potentially impacting the comprehensiveness of the models.

It is also important to note that our models did not include interaction effects of mutations with treatment and so did not explicitly model predictive biomarkers^36^. For example, in the case of NSCLC, ALK mutations were shown to be protective, likely due to the availability of ALK inhibitors as approved treatments. The absence of such interaction effects in our models limits the interpretation of the predictive value of specific mutations in the context of treatment response, and this is explored in other studies^37^.

Despite these limitations, our study offers a comprehensive, large-scale, and data-driven assessment of cancer that may be valuable for hypothesis generation, prognostic modeling, and risk stratification in oncology.

## Materials & Methods

### Data

Patient-level data and outcomes were derived from the pan-tumor CGDB offered jointly by Flatiron Health and Foundation Medicine. The CGDB is a US nationwide, longitudinal, de-identified oncology database that combines real-world, patient-level clinical data and outcomes with patient-level genomic data from over 280 US cancer clinics (approximately 800 sites of care). Comprehensive genomic profiling of >300 cancer-related genes on Foundation Medicine next-generation sequencing tests (including both current solid and liquid assays and legacy assays: FoundationOne CDx, FoundationOne Liquid CDx, FoundationOne Heme, FoundationOne, FoundationOne Liquid, FoundationACT) were linked to Flatiron EHR patient data via de-identified, deterministic matching^38–40^. To date, over 400,000 samples have been sequenced from patients across the US. The data are de-identified and subject to obligations to prevent reidentification and protect patient confidentiality. Altogether, the CGDB contains a rich set of thousands of potentially important prognostic factors for survival, including demographic characteristics, treatment regimens, disease and diagnosis profiles, mutational status of cancer-related genes, and longitudinal records of laboratory tests.

Patients from 16 cancer cohorts with a recorded oncology clinician-defined, rule-based, first line of therapy (1L) between January 1, 2011 through June 30, 2020 were pooled into a single, pan-cancer cohort containing 28,079 patients. The pan-cancer cohort comprised the following 16 cancer types: breast, chronic lymphocytic leukemia (CLL), colorectal, diffuse large B-cell lymphoma (DLBCL), gastric, head and neck, hepatocellular carcinoma (HCC), melanoma, multiple myeloma, non-small cell lung cancer (NSCLC), ovarian, pancreatic, prostate, renal, smal-cell lung cancer (SCLC), and urothelial. Patients were split into a train set (80%) and test set (20%) using stratified random sampling by cancer type.

### Feature engineering and preprocessing

All available data from the CGDB were used to derive a suite of features for each patient corresponding to 5 data modalities: clinical/demographic, laboratory/vital signs, treatment, cancer-specific (these 4 collectively referred to as “clinical”), and genomic. These features are summarized at a high level in Table S1. Features were eliminated in a data-driven way if they were zero-variance, near-zero-variance (dummy variables with fewer than 20 counts), or missing in over 30% of the pan-cancer cohort. Multiple imputation by chained equations (MICE) was used to impute clinical features and a k-nearest neighbors (kNN) approach^41^ was used to impute genomic features incorporating 2,566 samples from The Cancer Genome Atlas (TCGA) that had information on mutations of all three relevant types available: short variants (SNVs, indels), copy number alterations (CN), and rearrangements (fusions, RE)^42^. Since the TCGA data were derived from whole genome sequencing, we filtered the data to only those genes measured on Foundation Medicine panels. All imputation was performed in the train set separately from the test set to generate m=5 imputed datasets. Because of complex pooling, the results are presented for the first imputed dataset (m=1) and results were subjectively similar across imputed datasets. Specific feature engineering efforts are described below.

Clinical-demographic information included information on patient age, gender, race, smoking status, body mass index (BMI), cancer type, cancer stage at diagnosis, advanced or metastatic status of the cancer at baseline, Eastern Cooperative Oncology Group (ECOG) Performance Status, and a composite measure of comorbidity (the Elixhauser comorbidity index^43,44^) derived from structured EHR diagnosis code data. Treatment was represented in the form of indicators for the unique drug category (e.g. chemotherapy, immunotherapy, targeted/biologic, targeted/nonbiologic) received during the first line of therapy (1L). The number of unique drugs received in 1L, year of frontline therapy, time from diagnosis to first treatment, and treatment at an academic center (vs. community center) were also included.

Time series summaries of over 100 longitudinal laboratory tests and vital signs were computed within 2 time windows prior to the patient’s first line of therapy initiation date: 60 days (∼2 months) and 720 days (∼2 years). The following metrics were computed within each window: mean, median, variance, max, min, approximate entropy, difference between the last 2 values, slope of the last 2 values, total number of tests, ratio of number of tests to the available window of data observed for each patient, and, for lab tests, the proportion of labs that were abnormal. For comparability across patients and testing devices, lab values were normalized to their upper and lower limits of normal. Clinical input was obtained to assign thresholds of plausible lab and vital sign values; outlying values were set to missing and imputed.

Genomic features were generated from a single specimen per patient, choosing the specimen collected closest to index date if multiple were available. Binary indicator variables were populated from the mutations assessed by the specimen’s Foundation Medicine panel, coding each gene’s short variant, copy number, and rearrangement status separately. Gene-variant combinations that were not measured on a panel were coded as “NA” and were later imputed in the k nearest neighbors step. To summarize the alterations on a pathway level, we used a literature-derived list of gene sets and coded a pathway as impacted if any of its constituent genes had a reported mutation. Another feature type that introduced external information was the node2vec derived values, which were derived by averaging the node2vec^45^ embeddings vector of all affected genes in a specimen. The embeddings themselves were computed by running the reference implementation of the node2vec algorithm (https://github.com/aditya-grover/node2vec) with default settings using as input the human protein-protein interaction network available from the HitPredict effort^46,47^. For interpretability, <u>Fig. S12</u> presents the genomic variable contributions to the key protein interaction networks (“node2vec”) selected in the pan- and single-tumor models. A final set of features incorporating the observed mutation statuses and external data were the computed exposures to previously published mutational signatures^48^. These exposures were inferred via the SigsPack R package^49^. While the majority of specimen-derived features harnessed the Foundation Medicine mutation readout, a handful of standalone scores were also incorporated - tumor mutational burden, tumor purity, PDL1 status, estimated percentage of genome loss of heterozygosity, and microsatellite instability status.

Finally, cancer-specific features were obtained from records unique to each of the 16 cancer cohorts. These were included for their potential importance in predicting cancer-specific survival. Examples include the Gleason Score (a prognostic grading score for patients with prostate cancer), metastatic sites (for metastatic cancers such as breast cancer and non-small cell lung cancer), and transformation status (denoting transformation from follicular lymphoma to diffuse large b-cell lymphoma). These cancer-specific features exhibit “structured missingness^50^” in the pan-cancer cohort: available for one or few cancer types, and missing for the rest. As a result, they were not imputed outside of the relevant cancer cohort(s) and instead were set to 0.

Categorical variables were one-hot encoded with the reference level set to the majority level. All variables were normalized and outliers were truncated at +/- 3 z-scores for model stability.

### Model development

A penalized Cox proportional hazards model with lasso regularization was used to predict overall survival (OS) in the pan-cancer cohort using 2,059 (2,135 after one-hot encoding) CGDB-derived features. Survival time was calculated from 1L initiation date to death or last activity record in the EHR. Note that Flatiron EHR mortality records are validated against the National Death Index (NDI), widely considered a gold standard death dataset in the US, and shown to have high sensitivity, specificity, and date accuracy. A risk set adjustment was used to adjust for left truncation (see McGough *et al*.^51^ for a discussion on left truncation in this data source), and the model was adjusted for entry time to achieve quasi-independence between entry time and survival time^52^. Additionally, the model was adjusted for cancer type and compared to a stratified Cox model to account for potentially different baseline hazards by cancer type. Stratified Cox models were similar to but slightly outperformed by the non-stratified models (Fig. S13) and so are not described in the main text. Thus the main text describes pan-cancer models adjusted for cancer type. All models were fit using glmnet v. 4.0^53,54^ which handles left-truncated and right-censored survival data.

Patients were split into a train set (80%) and test set (20%) using stratified random sampling by cancer type. Five-fold cross-validation was used to tune the penalized model and the value of the lasso penalty, *λ*, that maximized the concordance index was selected to give the final model. Out-of-sample pan-cancer predictions were made on the withheld test set comprising (i) the overall pan-cancer cohort and (ii) each single-cancer cohort.

To compare predictions and insights gained from pan-cancer settings to those gained from single-cancer settings, a series of equivalent single-cancer models were developed dynamically using the original feature set. Feature normalization, detection and removal of zero- and near-zero-variance predictors, and truncation of outliers were performed separately in each single-cancer cohort, driven by the available data for that cancer type to simulate a real-world single-cancer setting.

Pan- and single-cancer models constructed on the full CGDB data were benchmarked against simpler models from clinical practice and the literature: (1) a benchmark model containing cancer type, age, gender, race, smoking status, cancer stage at diagnosis, baseline Eastern Cooperative Oncology Group (ECOG) Performance Status, time from diagnosis to initiation of 1L, and time from genomic test to initiation of 1L; and (2) a model adapted from ROPRO (Real wOrld PROgnostic score) by Becker et al^9^. These models are described in Table S2 and SI Materials & Methods.

### Model evaluation

Pan- and single-cancer models were evaluated using the out-of-sample concordance index (c-index) and integrated Brier score (IBS). Additionally, predicted risk scores were calculated for each patient as the exponential of the linear predictors from the penalized Cox model. Predicted risk scores were then used to stratify test set patients into high- and low-risk categories in each cancer cohort based on the median risk score of the training patients in the cohort.

Bias-corrected bootstrap percentile intervals^16^ were used to quantify uncertainty in model performance metrics using B=1,000 bootstraps of the train and test data.

### Software

All analyses were performed using R v. 4.1.1 (R Core Team 2021)^55^.

Data ingestion, manipulation, and preprocessing was performed using the R packages dplyr (v1.0.7)^56^, dbplyr (v2.1.1)^57^, rlang (v1.1.0)^58^, data.table (v1.14.0)^59^, tidyr (v1.1.3)^60^, stats^55^, purrr (v1.0.1)^61^, wrapr (v2.0.8)^62^, stringr (v1.4.0)^63^, hashmap (v0.2.2)^64^, pracma (v2.3.3)^65^, rsample (v0.1.0)^66^, fastDummies (v1.6.3)^67^, and coder (v0.13.5)^68^. Data imputation was performed using mice (v3.13.0)^69^ and impute (v1.65.0)^70^, and models were run using glmnet (v4.1-3)^53^, survival (v3.2-13)^71^, caret (v6.0-88)^72^, and LTRCforests (v0.5.5)^73,74^. Code parallelization and execution was performed using doParallel (v1.0.16)^75^, foreach (v1.5.1)^76^, doFuture (v0.12.0)^77^, parallel^55^, rngtools (v1.5)^78^, and doRNG (v1.8.2)^79^ and logged using logger (v0.2.1)^80^. Finally, figures were rendered using ggplot2 (v3.3.5)^81^.

## Data Availability

The data that support the findings of this study originated by Flatiron Health, Inc. and Foundation Medicine, Inc. Requests for data sharing by license or by permission for the specific purpose of replicating results in this manuscript can be submitted to.

## Acknowledgements

We thank F. Di Nucci, M. Hafner, S. Mahrus, and S. Maund for providing thoughtful feedback and suggestions for this study.

## Competing interests

The authors declare the following competing interests: R. Tibshirani and B. Narasimhan are paid consultants for Roche.

## Supplemental Information

**SI Tables & Figures:**

**Table S1.** Summary of features included in prognostic models.

**Table S2.** Summary of prognostic model feature sets.

**Figure S1-S5.** Pan-cancer and single-cancer risk stratification plots for all cancer types.

**Figure S6.** Integrated Brier Score (IBS) for pan-cancer and single-cancer (A) Benchmark, (B) ROPRO-like, and (C) Full models.

**Figures S7-9.** Top 25 clinico-genomic predictors in each single-cancer model.

**Figure S10.** Number of variables selected by each cancer model as a function of sample size.

**Figure S11.** Comparison of left-truncated right-censored forest (LTRCF) model performance for single-cancer and pan-cancer training cohorts with respect to the (A) c-index and (B) integrated brier score (IBS).

**Figure S12.** Top 15 variants associated with node2vec dimensions.

**Figure S13.** Comparison of model performance between the stratified and non-stratified Cox models with respect to (A) c-index and (B) integrated Brier score (IBS).

**SI Materials & Methods**

## Notes

### Funding Statement

R. Tibshirani and B. Narasimhan received funding as paid consultants of Roche.

### Author Declarations

IRB of Copernicus Group gave ethical approval on 07Feb2018 for the electronic health records database used in this analysis.

