## Supplemental Information for "Prognostic pan-cancer and single-cancer models: A large-scale analysis using a real-world clinico-genomic database"

### for

**SI Tables & Figures** **2**

Table S1. Summary of features included in prognostic models. 2

Table S2. Summary of prognostic model feature sets. 3

Figure S1-S5. Pan-cancer and single-cancer risk stratification plots for all cancer types. 4

Figure S6. Integrated Brier Score (IBS) for pan-cancer and single-cancer (A) Benchmark, (B) ROPRO-like, and (C) Full models. Lower IBS is indicative of better model calibration. Cancer types are arranged on the x-axis from largest to smallest sample size. 9

Figures S7-9. Top 25 clinico-genomic predictors in each single-cancer model. Predictors are ordered on the y-axis by descending coefficient (log hazard ratio). 10

Figure S10. Number of variables selected by each single-cancer model as a function of sample size. Cancer types are arranged on the x-axis from smallest to largest sample size. 13

Figure S11. Comparison of left-truncated right-censored forest (LTRCF) model performance for single-cancer and pan-cancer training cohorts with respect to the (A) c-index and (B) integrated brier score (IBS). Cancer types are arranged on the x-axis from largest to smallest sample size. 14

Figure S12. Top 10 variants associated with node2vec dimensions. Displayed node2vec dimensions are a subset of the full 128 that were chosen at least once by any model. Association was determined by the BIOT method. 15

Figure S13. Comparison of model performance between the stratified and non-stratified Cox models with respect to (A) c-index and (B) integrated Brier score (IBS). Cancer types are arranged on the x-axis from largest to smallest sample size. 16

**SI Materials & Methods** **17**

Adaptation of the ROPRO prognostic model 17

Investigation of alternative model fitting approaches that support interactions. 17

### SI Tables & Figures

**Table S1.** Summary of features included in prognostic models.

| Modality | # Features (raw) | # Features (one-hot encoded) | Type | Description |
|---|---|---|---|---|
| Clinical/demographic | 17 | 32 | Categorical, Continuous | Collection of baseline clinical/demographic information including: age, gender, race, BMI, ECOG, smoking status, cancer type, cancer stage at diagnosis, advanced or metastatic status at baseline, insurance type, socioeconomic status index, time from diagnosis to genomic test. |
| Labs and vital signs | 412 | 412 | Continuous | Time series summaries (e.g. median, min, max, proportion abnormal, slope, variance) of common labs and vital signs. |
| Treatment | 32 | 32 | Binary, Continuous | Indicators for each unique drug category received during frontline therapy and treatment at an academic center; continuous variable describing number of unique drugs received in frontline therapy, year of frontline therapy, time from diagnosis to first treatment. |
| Genomic | 1,511 | 1,513 | Categorical, Continuous | Binary alteration status (short variant "SV", copy number "CN", or rearrangement "RE") of genes; Raw variables are binary, while imputed ones are continuous due to the nature of KNN imputation (the resulting value reflects level of agreement between nearest neighbors). Variables consist of all combinations of HUGO gene symbol and variant type (SV, CN, RE) that were assayed by any Foundation Medicine test.<br>Pathway affected status - biological pathways are designated as affected if any of their constituent genes have any kind of alteration.<br>Node2Vec 128-dimensional embedding vector averages of all genes altered in a sample.<br>Non-alteration related features derived from or associated with Foundation Medicine data: tissue tumor mutational burden (tTMB), tumor purity (computationally derived), PDL1 status, estimated ancestry (fractional assignment to 5 superpopulations) |
| Cancer-specific | 87 | 146 | Categorical, Continuous | Prognostic factors relevant to one or more cancer types, including: sites of metastases, |

| | | | | |
|---|---|---|---|---|
| | | | | extranodal sites, disease subtypes, cancer-specific lab test results, disease-specific histologies. |
| **Total** | 2,059 | 2,135 | | |
| BMI: Body mass index; ECOG: Eastern Cooperative Oncology Group; HUGO: HUman Genome Organization. | | | | |

**Table S2.** Summary of prognostic model feature sets.

| Model | # Features | Description | Feature set |
|---|---|---|---|
| Benchmark | 9 | Collection of variables commonly collected in clinical practice | Cancer type, Age, Race, Gender, Smoking status, Baseline ECOG, Cancer stage at diagnosis, Time from diagnosis to frontline treatment, Time from diagnosis to genomic test |
| ROPRO-like | 29 | All variables included in the ROPRO model (Becker et al. 2020) with <= 30% missing in the database, plus time from diagnosis to genomic test (to adjust for delayed entry). | Cancer type, Age, Gender, Smoking status, Baseline ECOG, Cancer stage at diagnosis, BMI, Body weight, Body height, Heart rate, Hemoglobin, Systolic blood pressure, Diastolic blood pressure, Urea nitrogen, ALP, ALT, AST, Calcium, Creatinine, Total protein, Bilirubin, Albumin, Hematocrit, Glucose, Platelet count, Lymphocyte count, Monocyte count, Neutrophil count, Time from diagnosis to genomic test |
| Full | 2,059 | All variables derived from the clinico-genomic database. | All clinical and genomic predictors |

**Figure S1-S5.** Pan-cancer and single-cancer risk stratification plots for all cancer types.

**Figure S1.** Pan-cancer and single-cancer risk stratification plots for Non-Small Cell Lung, Colorectal, and Breast cancers.

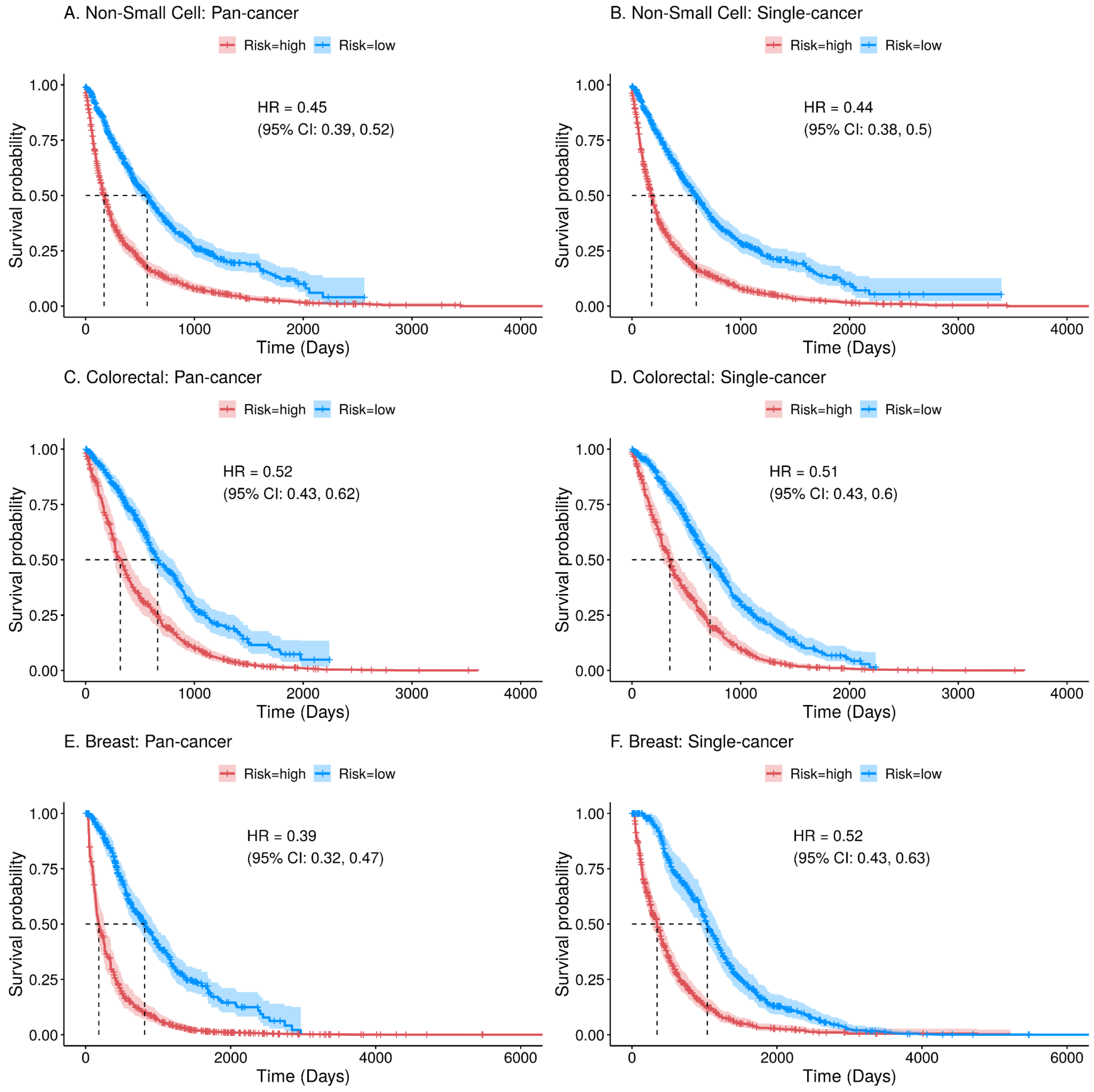

**Figure S2.** Pan-cancer and single-cancer risk stratification plots for Ovarian, Pancreatic, and Gastric cancers.

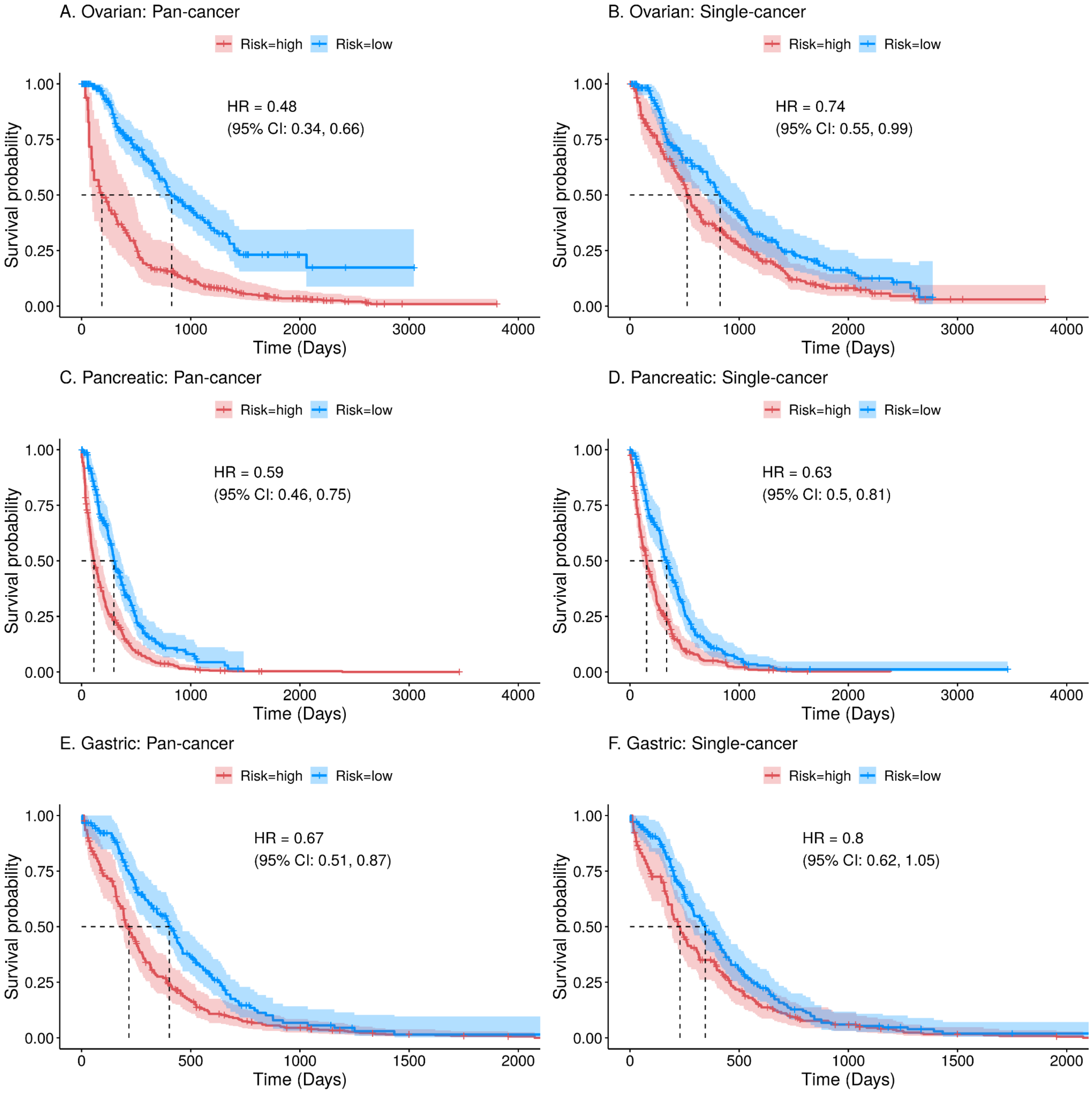

**Figure S3.** Pan-cancer and single-cancer risk stratification plots for Prostate, Urothelial, and Melanoma cancers.

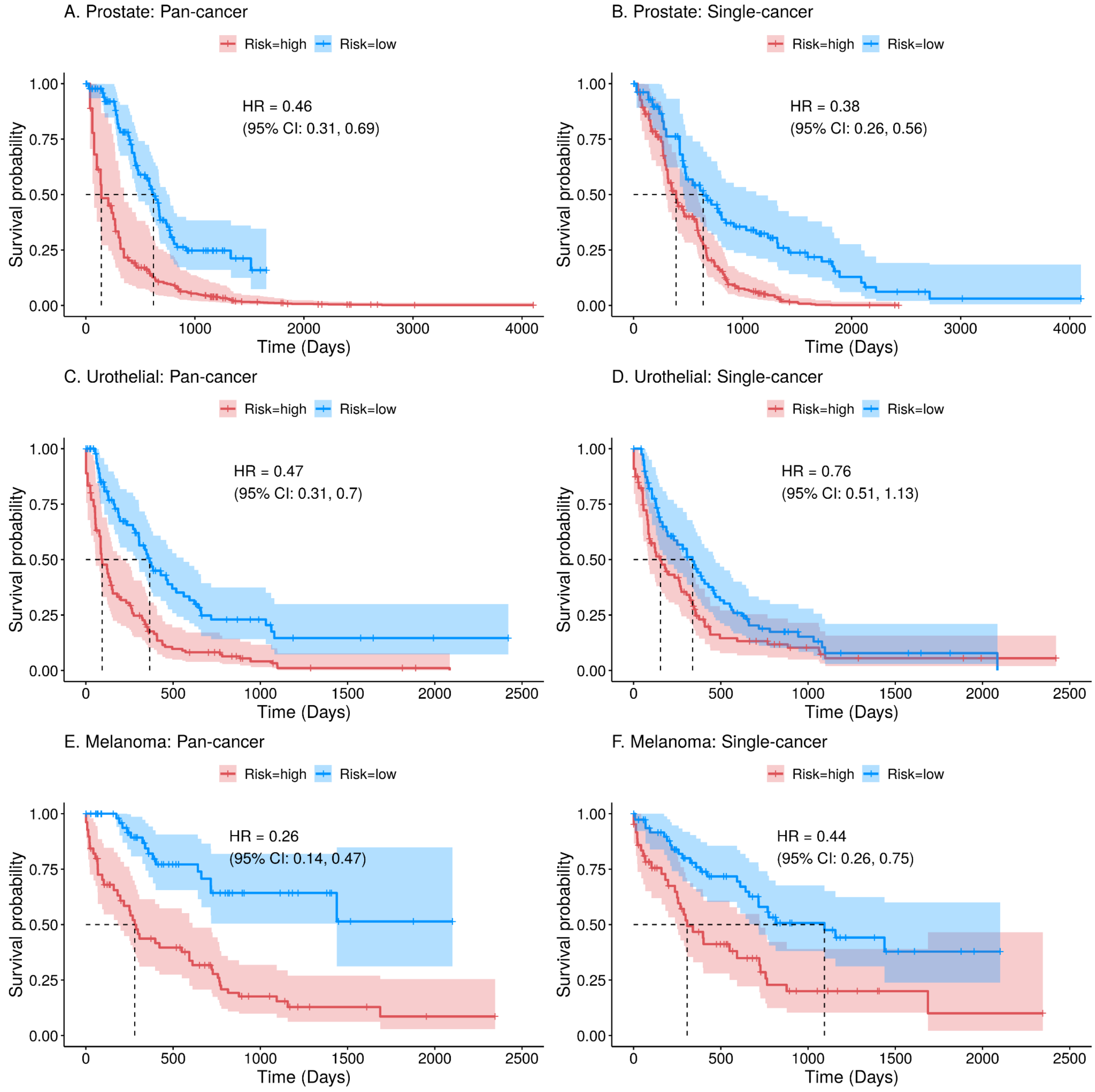

**Figure S4.** Pan-cancer and single-cancer risk stratification plots for Renal, Small Cell, and Head and Neck cancers.

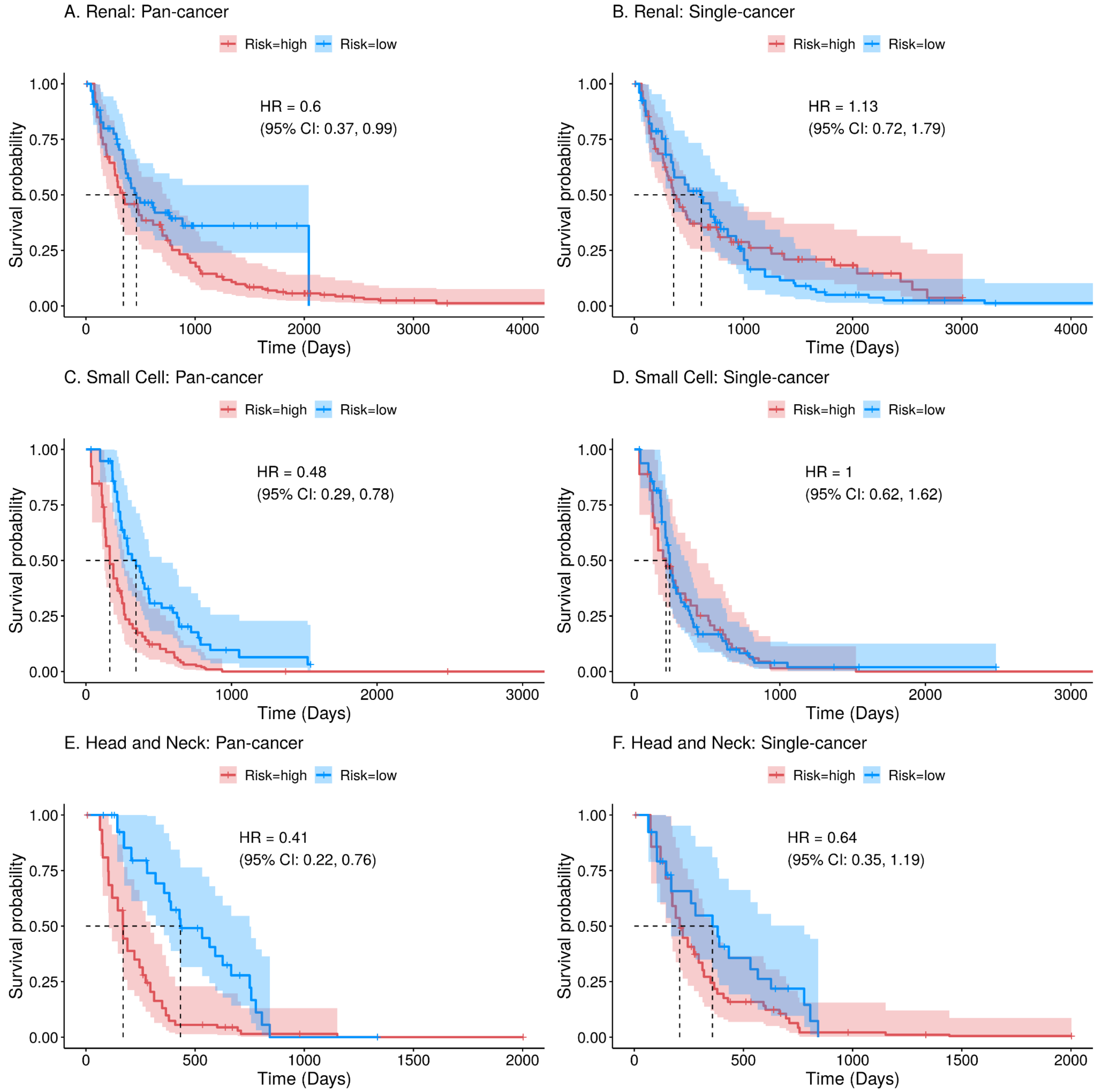

**Figure S5.** Pan-cancer and single-cancer risk stratification plots for Multiple Myeloma, Hepatocellular Carcinoma, DLBCL, and CLL.

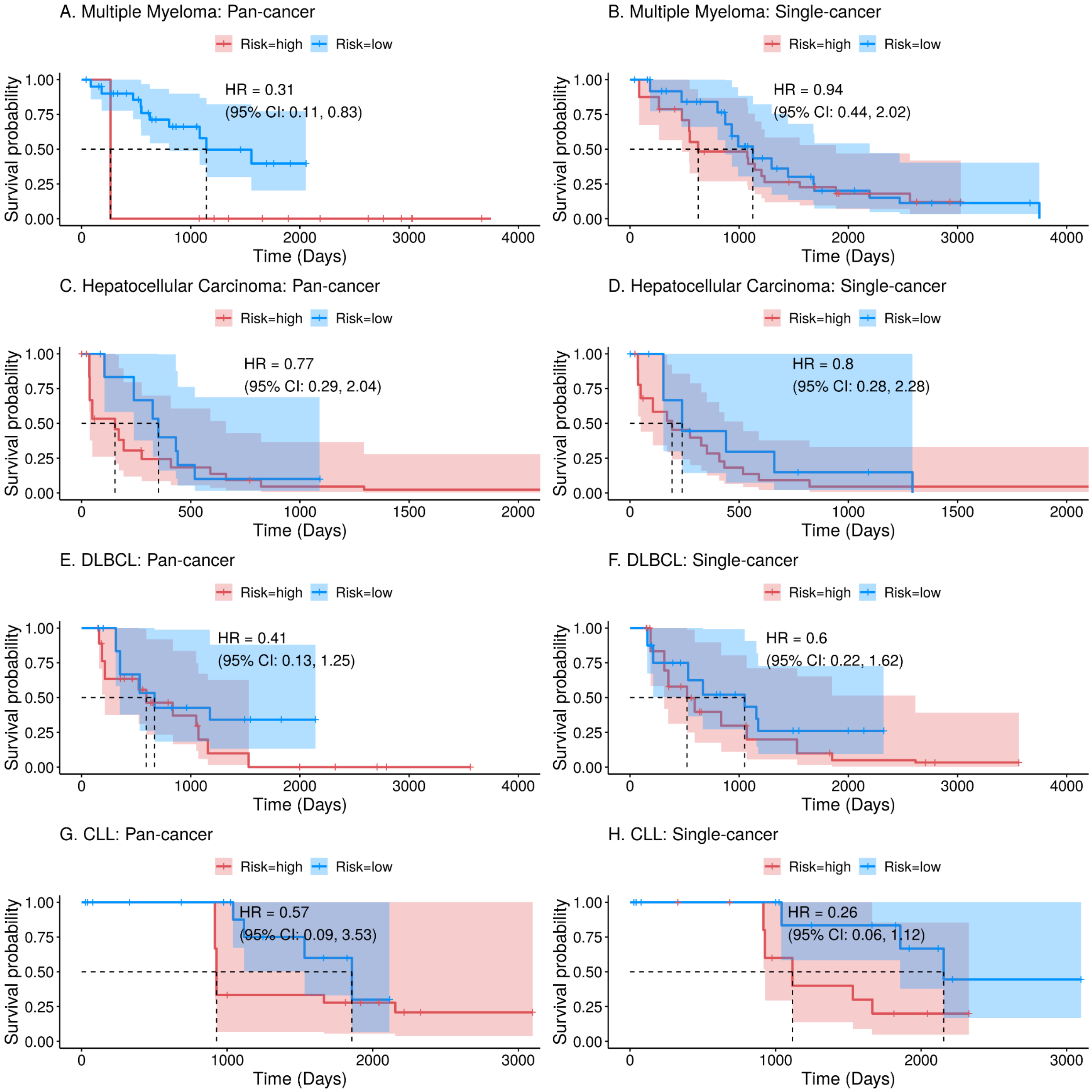

**Figure S6.** Integrated Brier Score (IBS) for pan-cancer and single-cancer (A) Benchmark, (B) ROPRO-like, and (C) Full models. Lower IBS is indicative of better model calibration. Cancer types are arranged on the x-axis from largest to smallest sample size.

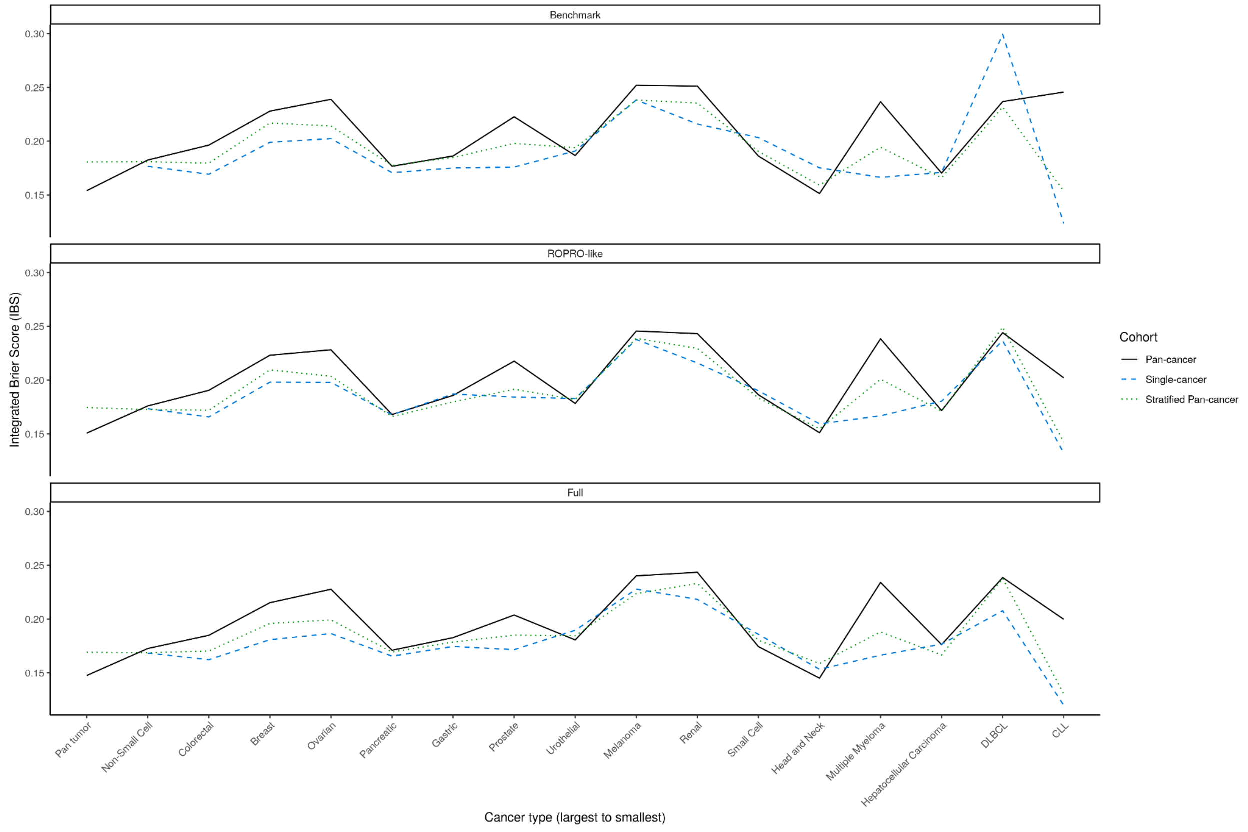

**Figures S7-9.** Top 25 clinico-genomic predictors in each single-cancer model. Predictors are ordered on the y-axis by descending coefficient (log hazard ratio).

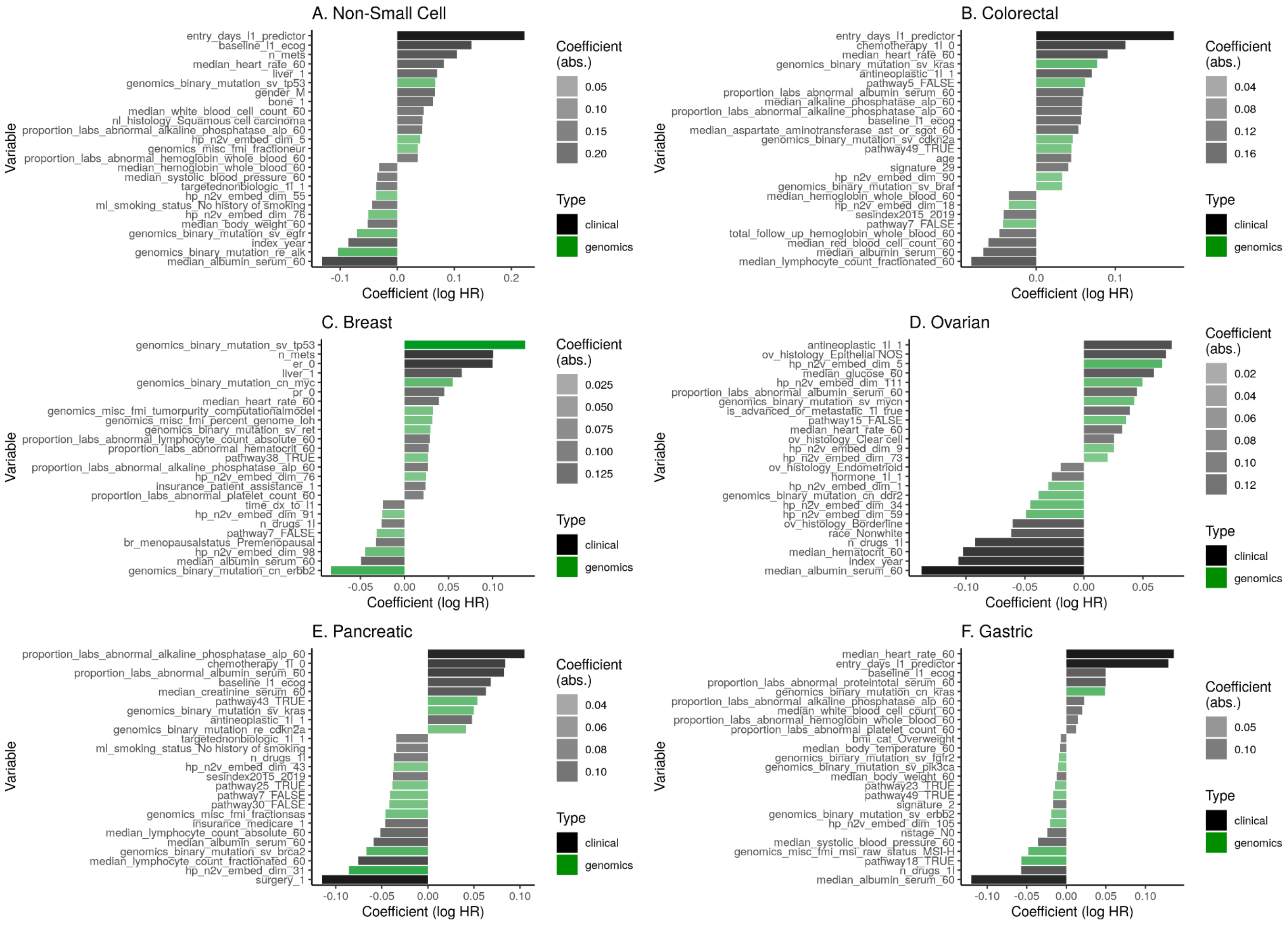

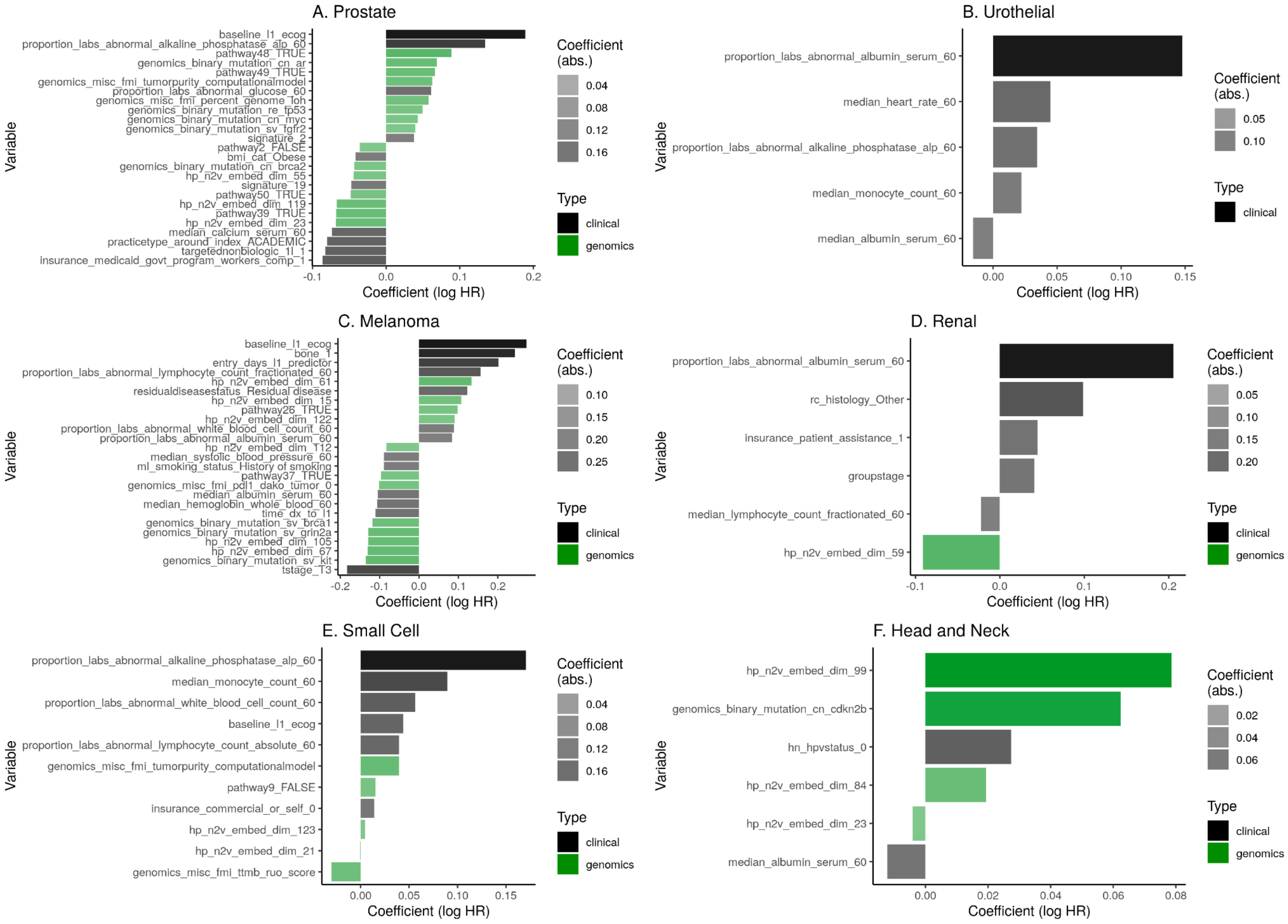
A. Prostate
B. Urothelial
C. Melanoma
D. Renal
E. Small Cell
F. Head and Neck
Variable
Coefficient (log HR)
Coefficient (abs.)
Type
clinical
genomics

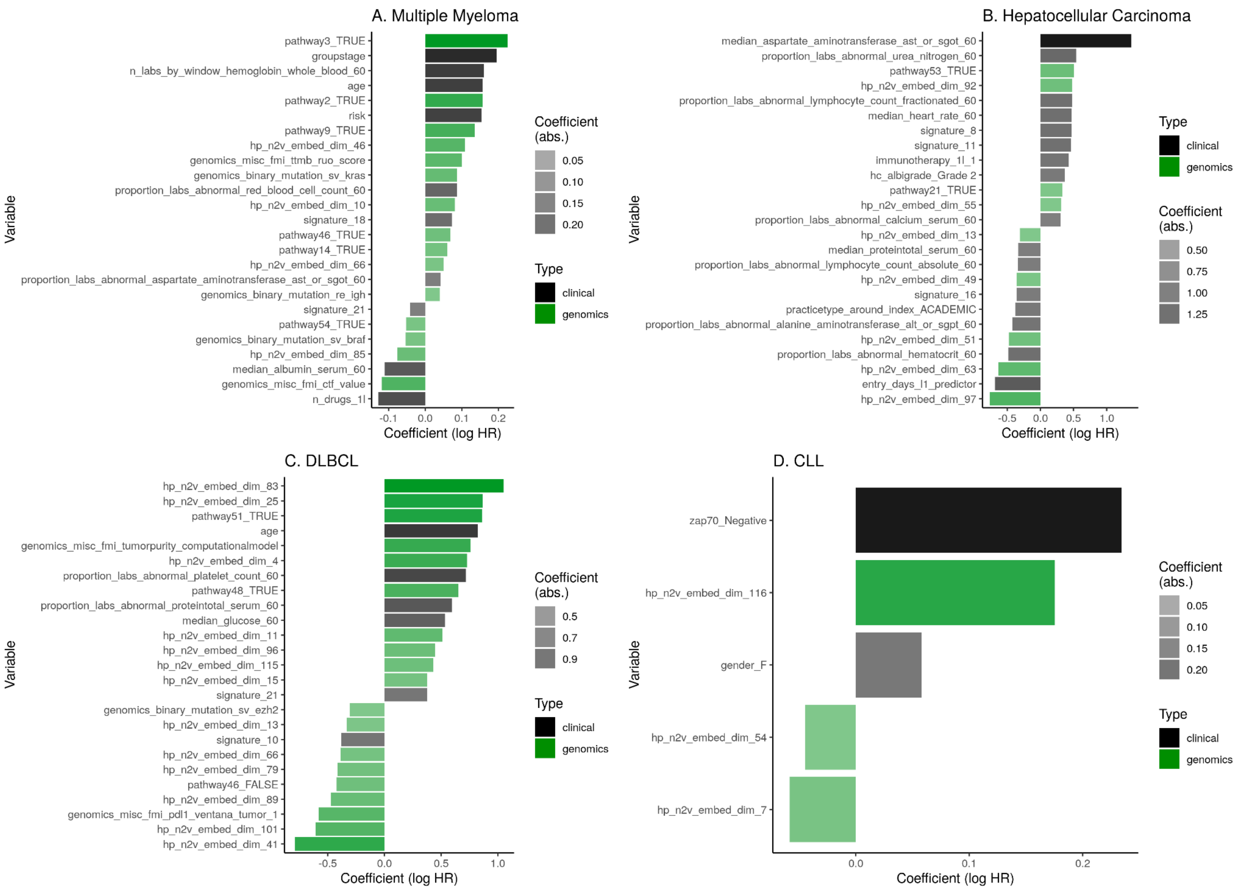
A. Multiple Myeloma
pathway3_TRUE
groupstage
n_labs_by_window_hemoglobin_whole_blood_60
age
pathway2_TRUE
risk
pathway9_TRUE
hp_n2v_embed_dim_46
genomics_misc_fmi_ttmb_ruo_score
genomics_binary_mutation_sv_kras
proportion_labs_abnormal_red_blood_cell_count_60
hp_n2v_embed_dim_10
signature_18
pathway46_TRUE
pathway14_TRUE
hp_n2v_embed_dim_66
proportion_labs_abnormal_aspartate_aminotransferase_ast_or_sgot_60
genomics_binary_mutation_re_igh
signature_21
pathway54_TRUE
genomics_binary_mutation_sv_braf
hp_n2v_embed_dim_85
median_albumin_serum_60
genomics_misc_fmi_ctf_value
n_drugs_1l
-0.1
0.0
0.1
0.2
Coefficient (log HR)
Variable
Coefficient (abs.)
0.05
0.10
0.15
0.20
Type
clinical
genomics
B. Hepatocellular Carcinoma
median_aspartate_aminotransferase_ast_or_sgot_60
proportion_labs_abnormal_urea_nitrogen_60
pathway53_TRUE
hp_n2v_embed_dim_92
proportion_labs_abnormal_lymphocyte_count_fractionated_60
median_heart_rate_60
signature_8
signature_11
immunotherapy_1l_1
hc_albigrade_Grade 2
pathway21_TRUE
hp_n2v_embed_dim_55
proportion_labs_abnormal_calcium_serum_60
hp_n2v_embed_dim_13
median_proteintotal_serum_60
proportion_labs_abnormal_lymphocyte_count_absolute_60
hp_n2v_embed_dim_49
signature_16
practicetype_around_index_ACADEMIC
proportion_labs_abnormal_alanine_aminotransferase_alt_or_sgpt_60
hp_n2v_embed_dim_51
proportion_labs_abnormal_hematocrit_60
hp_n2v_embed_dim_63
entry_days_l1_predictor
hp_n2v_embed_dim_97
-0.5
0.0
0.5
1.0
Coefficient (log HR)
Variable
Type
clinical
genomics
Coefficient (abs.)
0.50
0.75
1.00
1.25
C. DLBCL
hp_n2v_embed_dim_83
hp_n2v_embed_dim_25
pathway51_TRUE
age
genomics_misc_fmi_tumorpurity_computationalmodel
hp_n2v_embed_dim_4
proportion_labs_abnormal_platelet_count_60
pathway48_TRUE
proportion_labs_abnormal_proteintotal_serum_60
median_glucose_60
hp_n2v_embed_dim_11
hp_n2v_embed_dim_96
hp_n2v_embed_dim_115
hp_n2v_embed_dim_15
signature_21
genomics_binary_mutation_sv_ezh2
hp_n2v_embed_dim_13
signature_10
hp_n2v_embed_dim_66
hp_n2v_embed_dim_79
pathway46_FALSE
hp_n2v_embed_dim_89
genomics_misc_fmi_pdl1_ventana_tumor_1
hp_n2v_embed_dim_101
hp_n2v_embed_dim_41
-0.5
0.0
0.5
1.0
Coefficient (log HR)
Variable
Coefficient (abs.)
0.5
0.7
0.9
Type
clinical
genomics
D. CLL
zap70_Negative
hp_n2v_embed_dim_116
gender_F
hp_n2v_embed_dim_54
hp_n2v_embed_dim_7
0.0
0.1
0.2
Coefficient (log HR)
Variable
Coefficient (abs.)
0.05
0.10
0.15
0.20
Type
clinical
genomics

**Figure S10.** Number of variables selected by each single-cancer “full” model as a function of sample size. Cancer types are arranged on the x-axis from smallest to largest sample size.

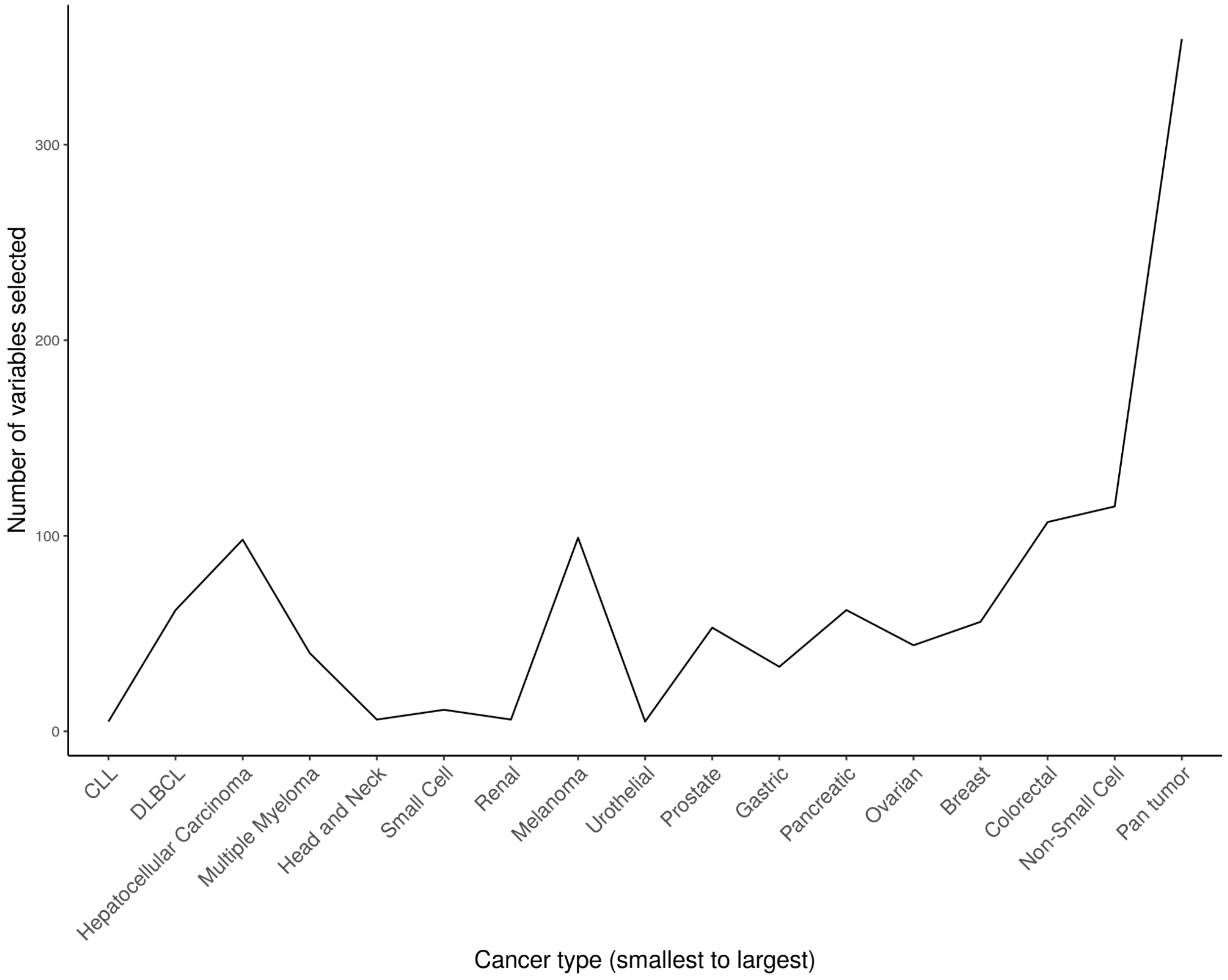

**Figure S11.** Comparison of left-truncated right-censored forest (LTRCF) model performance for single-cancer and pan-cancer training cohorts with respect to the (A) c-index and (B) integrated brier score (IBS). Cancer types are arranged on the x-axis from largest to smallest sample size.

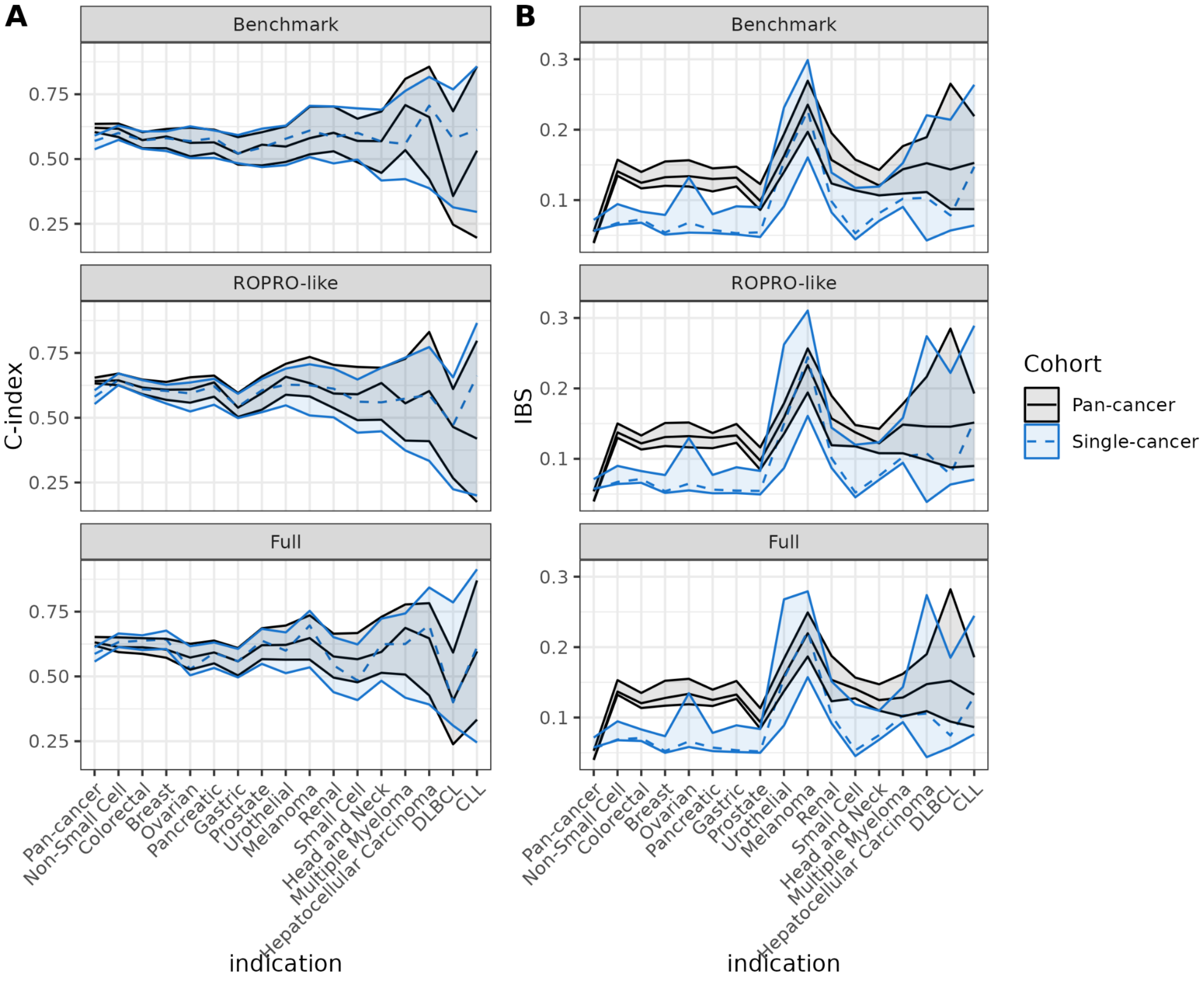

**Figure S12. Top 10 variants associated with node2vec dimensions**. Displayed node2vec dimensions are a subset of the full 128 that were chosen at least once by any model. Association was determined by the BIOT method.

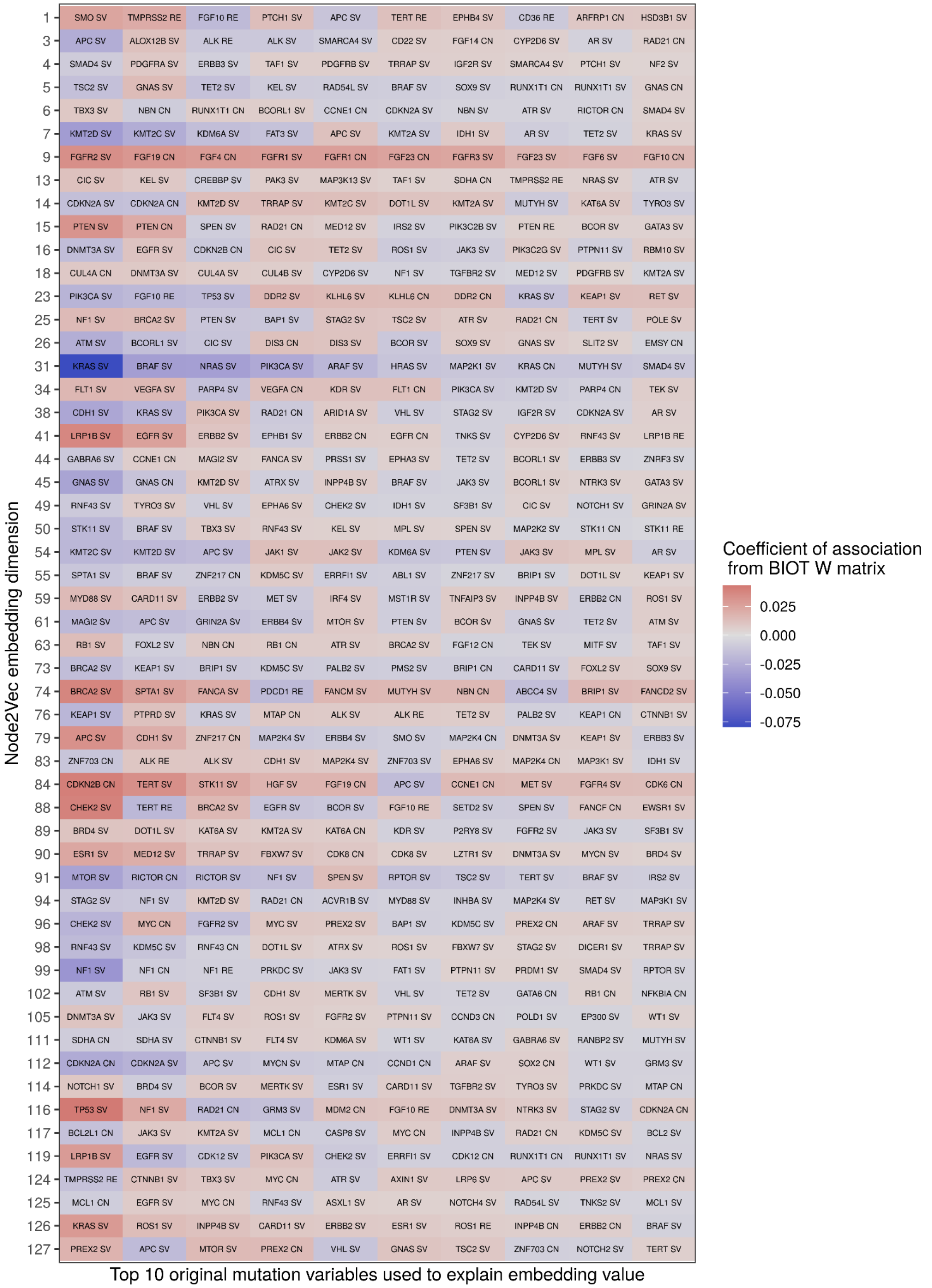

**Figure S13.** Comparison of model performance between the stratified and non-stratified Cox models with respect to (A) c-index and (B) integrated Brier score (IBS). Cancer types are arranged on the x-axis from largest to smallest sample size.

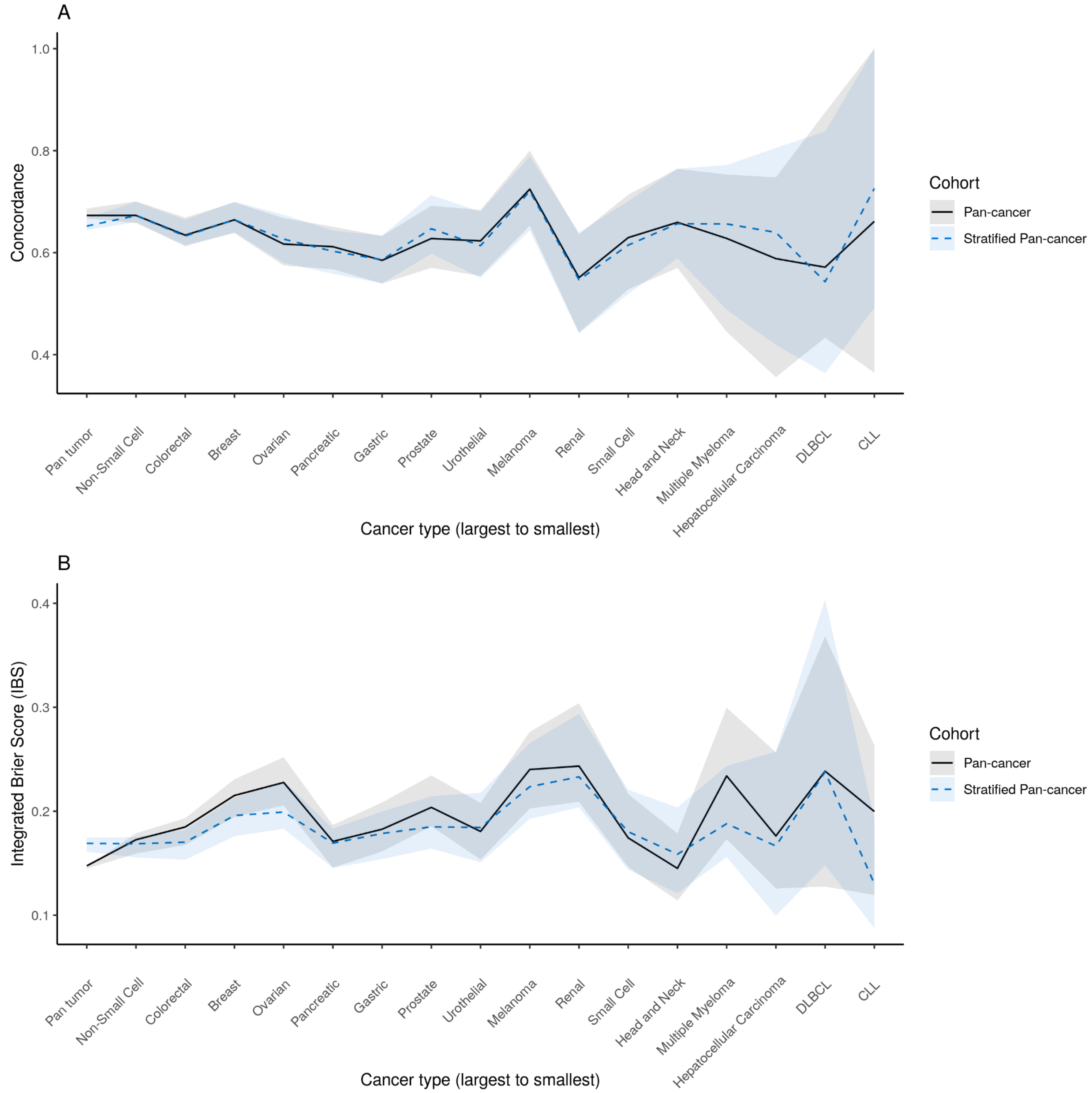

### SI Materials & Methods

#### *Adaptation of the ROPRO prognostic model*

The ROPRO (Real wOrld PROgnostic score) was developed by by Becker et al[9] and is composed of 27 clinical and demographic variables derived from the Flatiron Health electronic health record de-identified database.

We developed a “ROPRO-like” model, intended to adapt the ROPRO model using the available data in the clinico-genomic database (Table S2). However, our approach deviates in several ways:

1. To remain consistent with the rest of our benchmark models, we allowed missingness in the selected variables of up to 30% whereas the ROPRO model allowed missingness up to 75%. As a result, our “ROPRO-like” model excludes several ROPRO variables with high rates of missingness including lactate dehydrogenase (LDH), chloride, oxygen, and eosinophils.
2. Where variables like “AST-to-ALT ratio” were not available, we included the individual laboratory components, AST and ALT.
3. The ROPRO model imputed missing data using a tree-based approach, whereas we used multiple imputation to remain consistent across our benchmark models.

#### *Investigation of alternative model fitting approaches that support interactions.*

The left-truncated right censored forests (LTRCF) R package[73,74] was used within the same training and testing setup described for the Cox PH and Cox lasso models in the main results. The ‘mtry’ parameter was tuned via the LTRCforests::tune.ltrcrrf function (all arguments to which were set to their default values except time.eval, which was a grid from 0 to max observed time with granularity of 30 days. Default parameters: starting mtry = sqrt(number of variables), stepFactor = 2, ntreeTry = 100, bootstrap = “by.root”, samptype=”swor”, sampfrac=0.632, nsplit = 10, nodesizeTry = max of either sqrt(number of observations) or 15. Values of the linear predictor were obtained from the resulting fit using the LTRCforests:::predict.ltrcrfsrc method, and survival probabilities were calculated with the LTRCforests::predictProb method. These values were then used to calculate the final performance metrics: the concordance index and integrated Brier score.
